# Late diagnosis of Marfan syndrome is associated with unplanned aortic surgery and cardiovascular death

**DOI:** 10.1101/2024.04.22.24305998

**Authors:** Jason Claus, Lauritz Schoof, Thomas S. Mir, Anna Lena Kammal, Gerhard Schön, Kerstin Kutsche, Christian-Alexander Behrendt, Klaus Kallenbach, Tilo Kölbel, Christian Kubisch, Till Joscha Demal, Johannes Petersen, Jens Brickwedel, Michael Hübler, Christian Detter, Paulus Kirchhof, Eike Sebastian Debus, Meike Rybczynski, Yskert von Kodolitsch

## Abstract

**Background:** Marfan syndrome (MFS) guidelines recommend optimal pharmacological therapy (OPT) and replacement of the ascending aorta (RAA) at 5.0cm diameters to prevent acute type A aortic dissection (ATAAD) and death. The effect of early MFS diagnosis and initiation of therapy on outcomes is not known.

**Objective:** To evaluate the effect of age at MFS diagnosis and therapy initiation on delayed RAA and death.

**Methods:** This retrospective observational cohort study with long-term follow-up included consecutive patients with MFS, pathogenic *FBN1* variant, and regular visits to a European Reference Network Center. We considered MFS diagnosis at age ≥21 years late, and OPT initiation at age <21 years early. Outcomes were delayed RAA with aneurysm diameter >5.0cm or ATAAD, and death from all causes. We used landmark design starting at age 21 years to determine associations with outcomes.

**Results:** The study group consisted of 288 patients (45.1% male), including 169 patients with late MFS diagnosis (58.7%) and 63 with early OPT (21.9%). During mean follow-up of 25±14.7 years, 78 patients had delayed RAA, with 42 operations for ATAAD and 36 for aneurysms ≥5.0cm. There were 33 deaths, including 11 deaths late after ATAAD. All deaths were cardiovascular. Late diagnosis, but not early OPT, showed univariate association with delayed RAA (P<0.001) and death (P=0.025). Multivariate Cox regression analysis confirmed late diagnosis as predictor of delayed RAA (hazard ratio (HR)=8.01; 95% confidence interval (95%CI) 2.52-25.45; P<0.001) and death (HR=4.68; 95%CI 1.17-18.80; P=0.029).

**Conclusions:** Late diagnosis of Marfan syndrome is associated with delayed surgery and death.

## Introduction

Marfan syndrome (MFS) is a pleiotropic autosomal dominant disease that is caused by pathogenic variants in the gene encoding fibrillin-1 (*FBN1*). Due to the weakening of cardiovascular tissue, patients develop aortic aneurysms with the risk of rupture, acute type A aortic dissection (ATAAD) and death. Guidelines recommend optimal pharmacological therapy (OPT) with beta-blockers or angiotensin receptor blockers, or their combination, to slow aortic root dilatation, and elective replacement of the ascending aortic (RAA) at 5.0cm diameters to prevent ATAAD and death ^1^. Timely diagnosis of MFS, early initiation of OPT, and planned replacement of an enlarged aorta may be able to prevent aortic aneurysms with risk of rupture, reduce the need for unplanned operations, and thereby prolong life ^2^. However, evidence regarding the timing of MFS diagnosis and early OPT initiation on outcomes is limited. To evaluate the impact of late MFS diagnosis and early OPT initiation on delayed RAA and premature death, we conducted a retrospective observational cohort study with long-term follow-up using landmark design. All-cause mortality served as the secondary outcome. We analyzed consecutive patients with MFS and regular visits to the German hub of the European Reference Network on Rare Multisystemic Vascular Diseases, Germany (VASCERN-GE).

## Methods

### Patients, diagnostic assessment, and follow-up

Consecutive patients with MFS seen at the Hamburg VASCERN-GE tertiary care center between January 2008 to December 2021 were enrolled in the program. All patients participated at least 3 months, with regular visits of at least 12 months’ intervals at the center. Patients were included if they had MFS with positive Ghent criteria including the presence of a likely pathogenic or pathogenic *FBN1* variant ^3^. We excluded patients with rarer forms of MFS, including so-called “neonatal MFS” and patients with MFS and pathogenic variants in other disease genes such as *TGFBR2* ^4^, because their natural disease course may differ from that of patients with typical MFS secondary to *FBN1* variants ^5^. Recent population-based studies from Denmark found a median age of 19 years at MFS diagnosis with only 5% risk for aortic events at age <21 years ^6^. Therefore, we considered MFS diagnosis late at ages ≥21 years, and OPT early with age <21 years. The primary outcome was delayed RAA, which we defined as RAA for aneurysms with diameters >5.0cm or for ATAAD. All study patients gave informed consent. This study was approved by the Ethics Committee of the Hamburg Medical Association in Hamburg, Germany, under registration number 2012-10518-BO-ff. The analysis adheres to the STROBE reporting criteria (Table S1 in the Supplemental Appendix)^7^.

To establish the first clinical diagnosis of MFS, we accepted MFS diagnoses from other institutions, and we noted the patient age at diagnosis according to medical records or, in the case of previously undiagnosed MFS, after we established the MFS diagnosis at our center. We confirmed the diagnosis of MFS in all patients according to the revised Ghent criteria ^4^: In the absence of a positive family history of MFS, MFS was present with aortic root dilatation in combination with ectopia lentis or a pathogenic *FBN1* variant or with a systemic Ghent score ≥7 points or with the combination of ectopia lentis with a pathogenic *FBN1* variant known to cause aortic dilatation. In the presence of a positive family history, MFS was present if ectopia lentis or a systemic Ghent score ≥7 points or aortic root dilatation was detected ^4^. A total of 321 patients met MFS criteria based on clinical criteria alone, and 20 patients met MFS criteria only in combination with clinical criteria and a likely pathogenic or pathogenic *FBN1* variant. Diagnostic confirmation of MFS required the presence of a likely pathogenic or pathogenic *FBN1* variant in all 321 patients. For genetic testing of *FBN1*, DNA was isolated from leukocytes by standard procedures. Until 2016, Sanger sequencing of the coding region and exon-intron boundaries identified the *FBN1* pathogenic variant. Since 2016, targeted next-generation sequencing panels replaced Sanger sequencing ^8^. We classified *FBN1* variants (mRNA reference sequence: NM_000138.5) as pathogenic or likely pathogenic (class IV-V) according to established standards ^3^.

### Outcomes

We assessed the duration from landmark age 21 years to most recent contact as the time of follow-up, and the duration from the age at first to the most recent visit at VASCERN-GE as time of care. Guidelines required at least annual appointments, where we adjusted intervals to 3 months or 6 months ^1^. Patients who failed to attend appointments received reminder calls and letters. Failure to attend regular appointments for ≥2 years was considered lost to follow-up, with documentation of clinical events and reasons for absence. To prevent ATAAD, guidelines recommend RAA for MFS at aortic diameters of 5.0cm ^1^. We considered timely RAA at diameters ≤5.0cm, and delayed RAA with one of the following criteria: First, RAA for aneurysms with diameters >5.0cm, because these diameters increase the risk of ATAAD requiring urgent RAA ^1^. Second, RAA for ATAAD requiring emergent RAA ^1^. ATAAD was present in the case of overt dissection or intramural hematoma involving the ascending portion of the aorta with matching symptoms ^1^. Finally, we considered all-cause mortality as death from any cause. Delayed RAA served as primary study outcome, and all-cause mortality as secondary outcome. Regardless of the center where surgery was performed, surgical techniques of RAA comprised isolated replacement of the supra-coronary part of the ascending aorta with an aortic tube graft ^9^, and complete replacement including the aortic root with a composite valve graft according to Bentall with biological or mechanical valve prostheses, or with aortic-valve-sparing procedures according to David or to Yacoub ^1^.

### Capture and completeness of events, certainty of evidence

To estimate the certainty of evidence, we presented data from all patients with MFS who were excluded because their age was <21 years, or who were lost to follow-up. To evaluate outcome bias, we examined all autopsies at the Institute of Forensic Medicine, University Medical Center, between January 2008 and December 2021 for fatal ATAAD with a pre-mortem diagnosis of MFS. To determine whether our cohort approximates the true number of MFS in the general population, we used the prevalence of MFS in the Danish population as a reference standard, the total number of 4,958,621 inhabitants in the Hamburg metropolitan region, and all 280 patients with MFS at VASCERN-GE in this region, including 217 study patients, 39 patients aged <21 years, 6 with neonatal MFS, 12 with negative and 6 without *FBN1* gene testing ^10, 11^. Finally, to validate the all-cause mortality of MFS, we used previously established routines to assess mortality of MFS and degenerative aortic disease from health insurance claims of 6.5 million insured patients of the health insurance company DAK-Gesundheit between 1/2008 and 12/2017 ^12^. For Kaplan-Meier curve analysis, we used ICD codes Q87.4 to identify 198 patients with MFS and ICD codes I70.0, I71, I74.0, or I74.1 to identify 46,533 patients with degenerative aortic disease. We performed a brief scoping review to qualify the nature of the evidence in the literature on the prognostic impact of time of diagnosis of MFS on aortic events and mortality, using the PRISMA extension for scoping review criteria (PRISMA-ScR; Table S2) ^13^. In our study limitations, we applied GRADE criteria for prognostic factors research to rate the certainty of evidence that a late diagnosis of MFS actually affects outcomes ^14^.

### Power calculation

To estimate study imprecision, we determined the minimum sample size of the study using a web-based calculator (https://riskcalc.org), setting the 2-sided significance level at 0.05, the study power (1-beta) at 0.8, the sampling ratio at 1.0, the probability of delayed RAA (primary endpoint) in the unexposed and exposed groups at 0.4 and 0.6, respectively, and the failure rate set at 20%.

### Statistical analysis

We used age at MFS diagnosis to distinguish two study groups according to early MFS diagnosis with age <21 years and late MFS diagnosis with age ≥21 years. We evaluated the following risk variables: Male sex, as this is associated with an increased risk of aortic events ^6^; systemic Ghent score ≥7 points, as this indicates a more severe MFS phenotype ^4^; early OPT with OPT initiation before the age of 21 years, as OPT before that age is considered most effective in slowing aortic growth rates ^2^; RAA before the age of 21 years, as RAA before that age is uncommon and indicates a more severe phenotype of MFS ^11^; sporadic occurrence of MFS with the presence of a *de novo FBN1* variant in an affected patient, as severe phenotypes are more likely with sporadic occurrence than with familial occurrence ^2^; genetic testing of index patients for disease-causing *FBN1* variants, as severe phenotypes are more likely in index patients than in relatives with cascade gene testing ^15^; residence in the Hamburg metropolitan area according to the geographic definitions of the 2011 census, as patients from this area more representative of the general German population than patients who traveled from other parts of Germany ^10^. Continuous data are presented as means ± standard deviations (SD), and categorical data are presented as counts and percentages, with the Kruskal-Wallis test or Fisher’s generalized exact method for comparisons. We used the Kaplan-Mayer method for bivariate analysis of time to event to represent the effects of exposure variables on outcomes, using log-rank to test for significant differences. We performed multivariate Cox regression analysis including all study covariates to determine whether late MFS diagnosis was an independent predictor of outcomes. There were no missing data in this study, all p-values were two-sided, and a p-value <0.05 was considered significant. Statistical software R version 4.3.2 was used for all statistical tests and plots.

## Results

### Population

A total of 365 patients fulfilling the Ghent criteria of MFS were potentially eligible for study inclusion. Of these, 6 patients were excluded because they had neonatal MFS, 12, because the *FBN1* gene testing was negative, and 6, because *FBN1* gene testing was not available. Of the remaining 341 patients, we excluded 53 patients because they did not reach landmark age 21 years. As a result, the final study group consisted of 288 patients, which exceeded the minimum total sample size of 268 patients required to avoid study imprecision. A screening of 279 autopsies with death by ATAAD in 162 men and 117 women at a mean age of 64±15 years did not identify patients with pre-mortem diagnosis of MFS (Figure S1). The 53 patients who were excluded from the study because they had not reached the landmark age of 21 years had lower mean age at diagnosis, as expected, lower mean age at first contact with VASCERN-GE, lower systemic score points, and lower frequency of genetic testing in the index patients than the study patients, and only two of them required RAA, and none experienced ATAAD or death (Table S3). Among the 26 patients who discontinued follow-up, the study variables were similar to all other study patients, and we identified specific reasons for discontinuation in 17 of them (65.4%; Table S4). Study groups

With a mean age of 28.5±15.9 years at MFS diagnosis (1-80 years), the study cohort included 169 patients with late (58.7%) and 119 patients with early MFS diagnosis (41.3%; Figure 1). The distribution of male sex, sporadic occurrence of MFS, and residence inside metropolitan area was comparable between both diagnosis groups. However, patients with late MFS diagnosis were older at first contact with VASCERN-GE, systemic Ghent score ≥7 points, OPT initiation and RAA before the age of 21 years, and relatives of index patients were less common, and genetic testing of index patients was more common than with early MFS diagnosis (Table 1).

**Figure 1.**
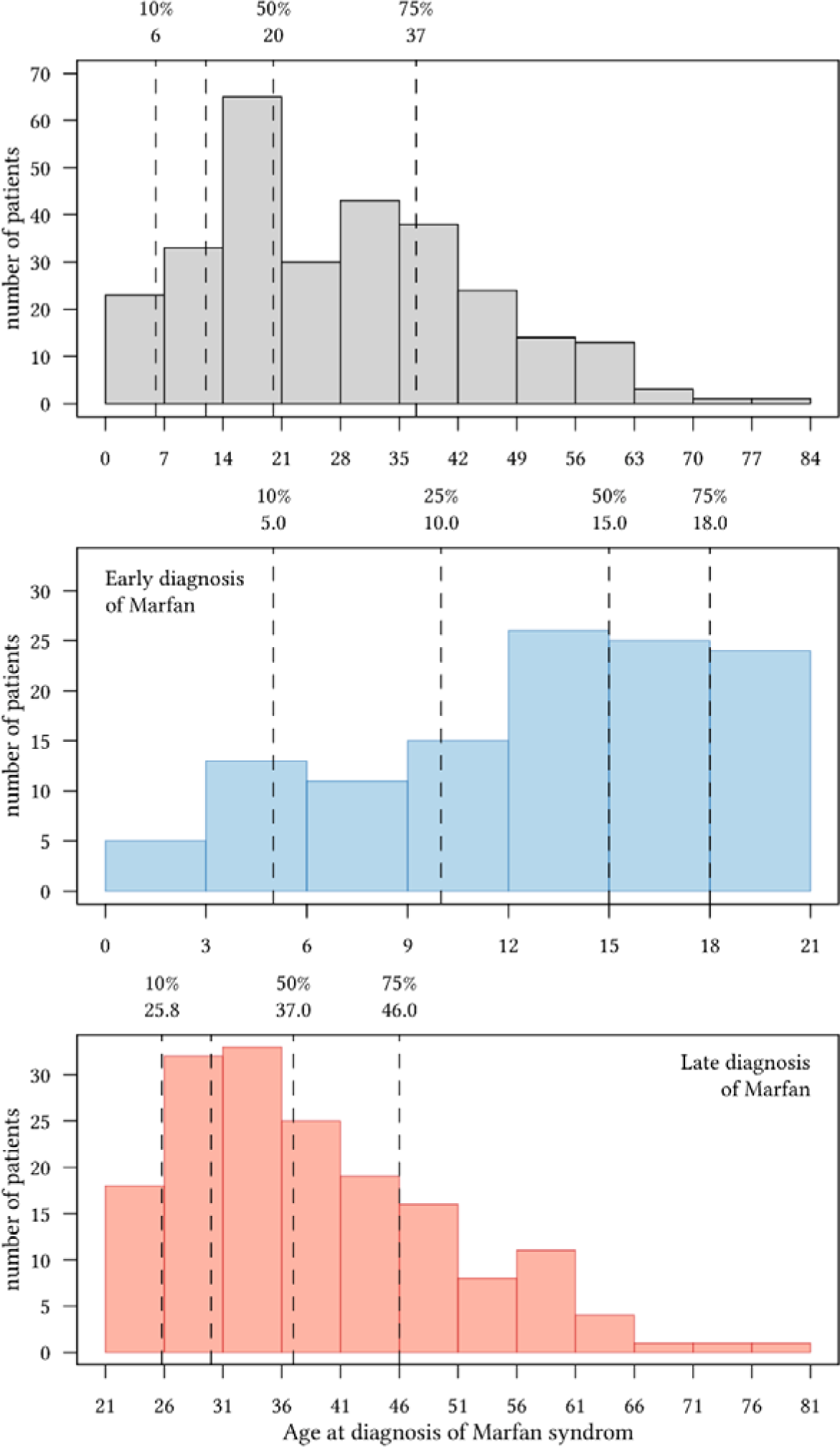
Diagnosis of Marfan syndrome according to age. Number of patients with Marfan syndrome by age at diagnosis during the study period 2008 to 2021. Dashed lines indicate the age when 10, 25, 50 and 75 % of patients are diagnosed.

**Table 1.**
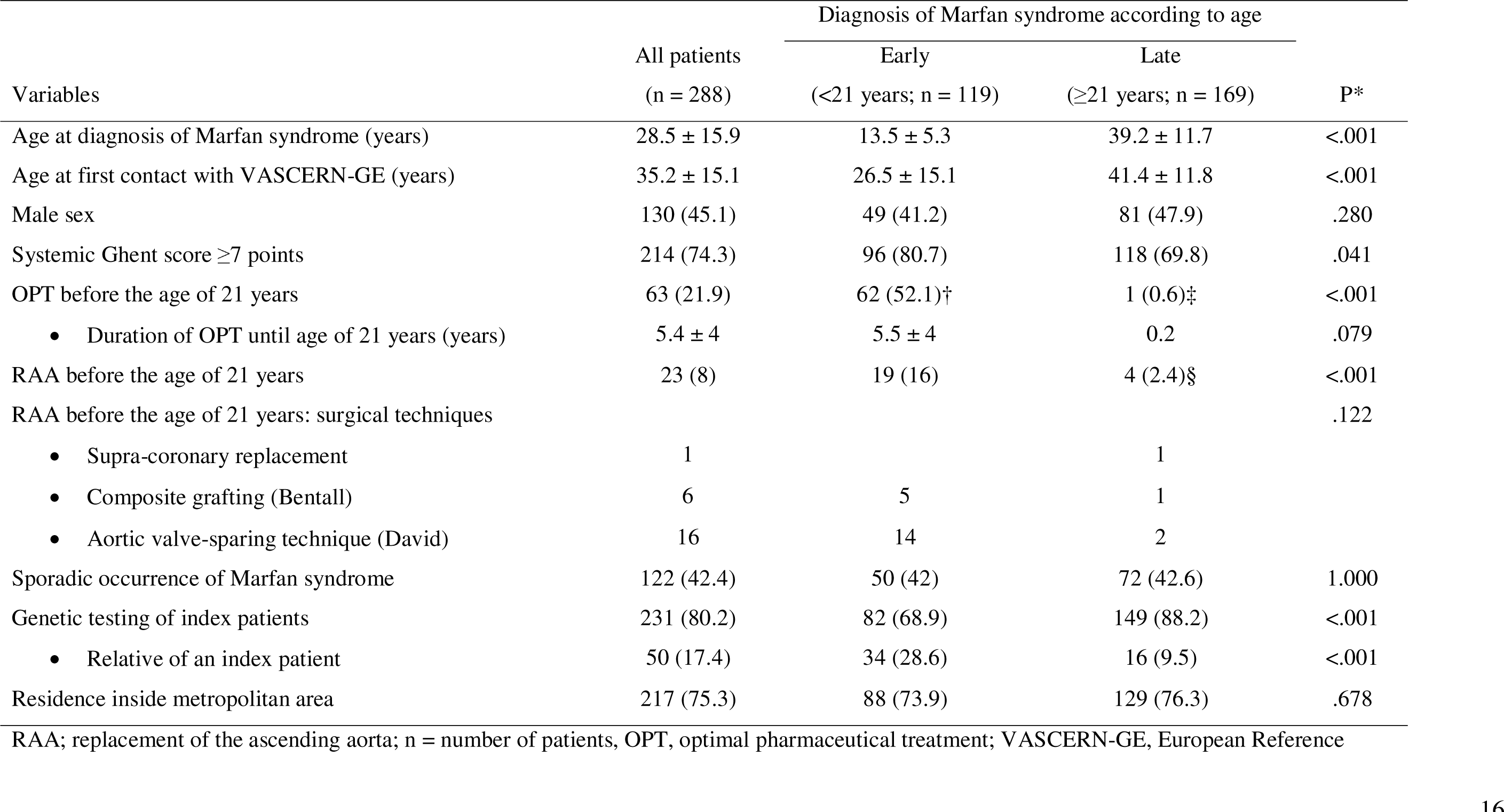
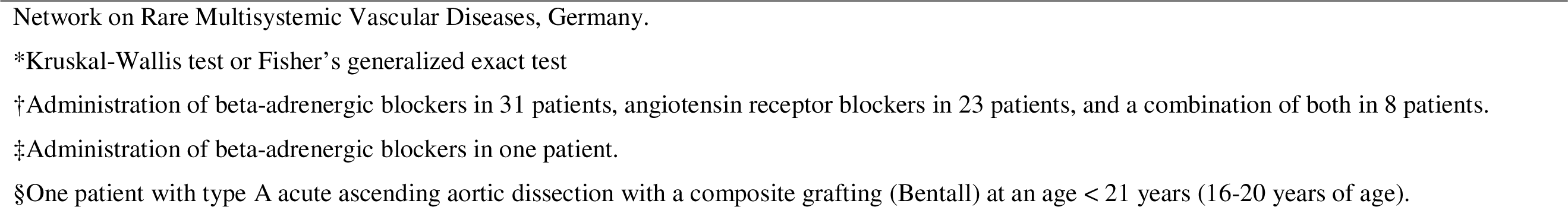
Study groups.

| Variables | All patients<br>(n = 288) | Diagnosis of Marfan syndrome according to age |  | P* |
| --- | --- | --- | --- | --- |
|  |  | Early<br>(<21 years; n = 119) | Late<br>(≥21 years; n = 169) |  |
| Age at diagnosis of Marfan syndrome (years) | 28.5 ± 15.9 | 13.5 ± 5.3 | 39.2 ± 11.7 | <.001 |
| Age at first contact with VASCERN-GE (years) | 35.2 ± 15.1 | 26.5 ± 15.1 | 41.4 ± 11.8 | <.001 |
| Male sex | 130 (45.1) | 49 (41.2) | 81 (47.9) | .280 |
| Systemic Ghent score ≥7 points | 214 (74.3) | 96 (80.7) | 118 (69.8) | .041 |
| OPT before the age of 21 years | 63 (21.9) | 62 (52.1)† | 1 (0.6)‡ | <.001 |
| • Duration of OPT until age of 21 years (years) | 5.4 ± 4 | 5.5 ± 4 | 0.2 | .079 |
| RAA before the age of 21 years | 23 (8) | 19 (16) | 4 (2.4)§ | <.001 |
| RAA before the age of 21 years: surgical techniques |  |  |  | .122 |
| • Supra-coronary replacement | 1 |  | 1 |  |
| • Composite grafting (Bentall) | 6 | 5 | 1 |  |
| • Aortic valve-sparing technique (David) | 16 | 14 | 2 |  |
| Sporadic occurrence of Marfan syndrome | 122 (42.4) | 50 (42) | 72 (42.6) | 1.000 |
| Genetic testing of index patients | 231 (80.2) | 82 (68.9) | 149 (88.2) | <.001 |
| • Relative of an index patient | 50 (17.4) | 34 (28.6) | 16 (9.5) | <.001 |
| Residence inside metropolitan area | 217 (75.3) | 88 (73.9) | 129 (76.3) | .678 |

RAA; replacement of the ascending aorta; n = number of patients, OPT, optimal pharmaceutical treatment; VASCERN-GE, European Reference
Network on Rare Multisystemic Vascular Diseases, Germany.
\*Kruskal-Wallis test or Fisher's generalized exact test
†Administration of beta-adrenergic blockers in 31 patients, angiotensin receptor blockers in 23 patients, and a combination of both in 8 patients.
‡Administration of beta-adrenergic blockers in one patient.
§One patient with type A acute ascending aortic dissection with a composite grafting (Bentall) at an age < 21 years (16-20 years of age).

### Outcomes

Follow-up time from the age 21 years to most recent contact with VASCERN-GE was longer (30.4±12.2 versus 17.4±14.6 years; P<0.001), time of care at VASCERN-GE was shorter (10.1±5.6 versus 12±5.5 years; P=0.010), but the number of transthoracic echocardiography examinations per year, and loss to follow-up was comparable in patients with late and early MFS diagnosis. Patients were more likely to experience primary (P<0.001) and secondary outcomes with late vs early MFS diagnosis (P<0.001). Patients underwent a total of 164 RAA procedures (56.9%). There were 23 RAA before the age of 21 years, but these were less frequent with late vs early MFS diagnosis (P<0.001; Table 2). There were 141 RAA at or after age 21 years. Of these, timely RAA only showed a trend towards differences (P=0.082), but delayed RAA for aneurysms >5.0cm or for ATAAD was more common with late vs early MFS diagnosis (P<0.001). All 42 RAA for ATAAD and all 11 RAA with isolated supra-coronary replacement were performed with late MFS diagnosis (Table 3).

**Table 2.**
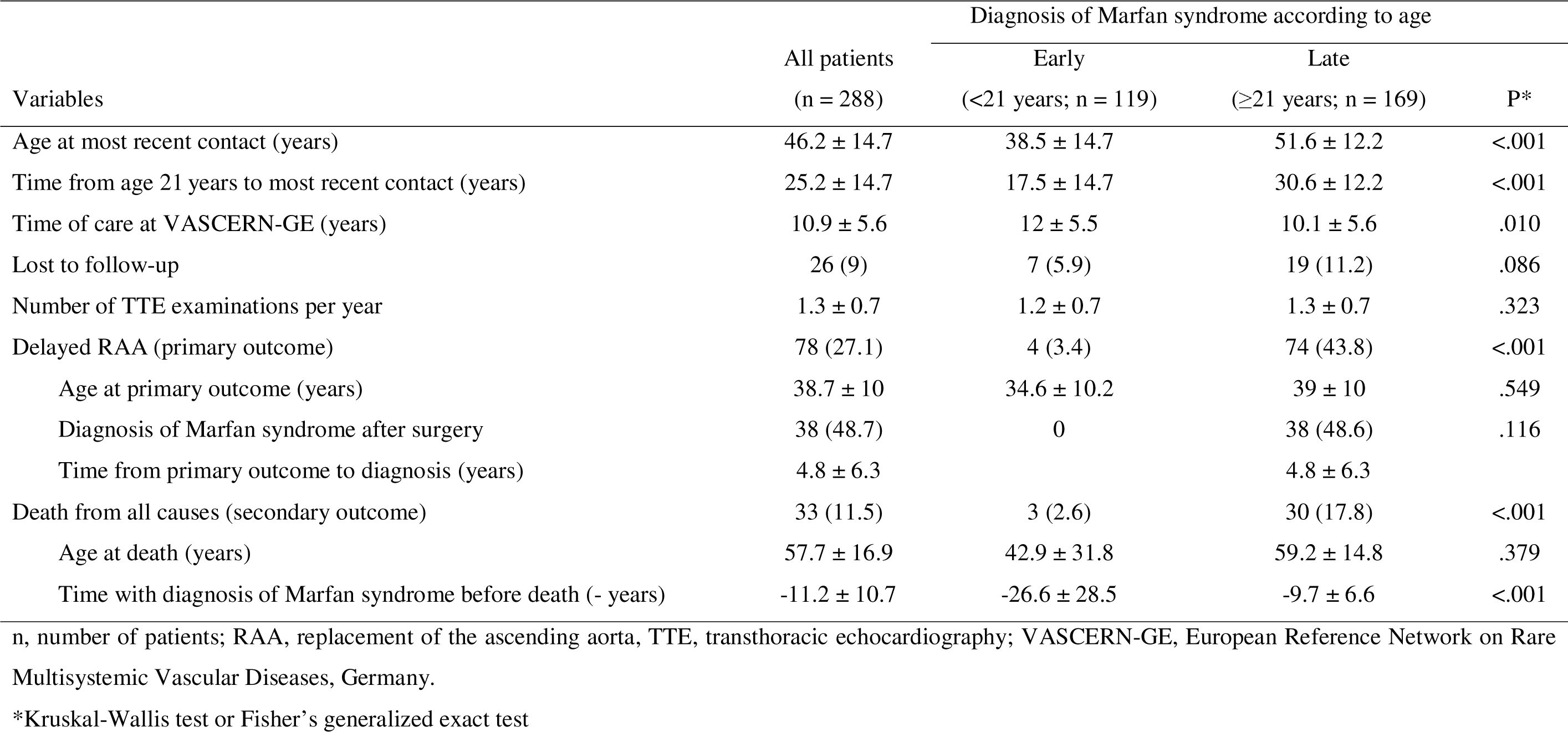
Outcomes.

**Table 3.**
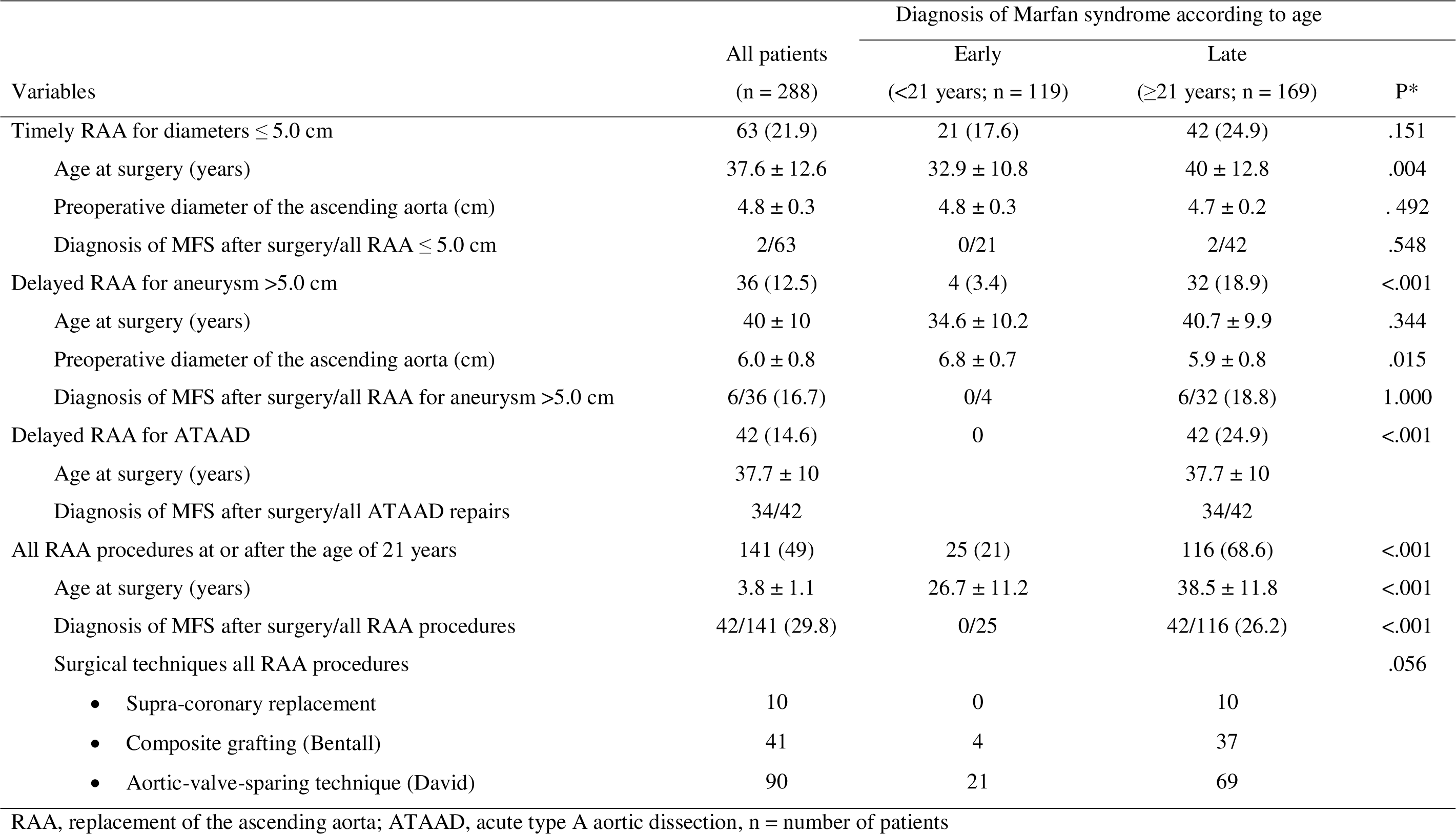
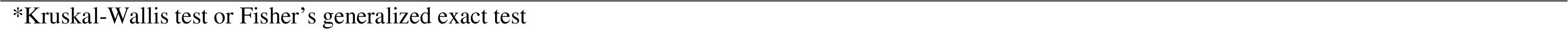
Repair of the ascending aorta at or after the age of 21 years.

| Variables | All patients<br>(n = 288) | Diagnosis of Marfan syndrome according to age |  | P* |
| --- | --- | --- | --- | --- |
|  |  | Early<br>(<21 years; n = 119) | Late<br>(≥21 years; n = 169) |  |
| Timely RAA for diameters ≤ 5.0 cm | 63 (21.9) | 21 (17.6) | 42 (24.9) | .151 |
| Age at surgery (years) | 37.6 ± 12.6 | 32.9 ± 10.8 | 40 ± 12.8 | .004 |
| Preoperative diameter of the ascending aorta (cm) | 4.8 ± 0.3 | 4.8 ± 0.3 | 4.7 ± 0.2 | .492 |
| Diagnosis of MFS after surgery/all RAA ≤ 5.0 cm | 2/63 | 0/21 | 2/42 | .548 |
| Delayed RAA for aneurysm >5.0 cm | 36 (12.5) | 4 (3.4) | 32 (18.9) | <.001 |
| Age at surgery (years) | 40 ± 10 | 34.6 ± 10.2 | 40.7 ± 9.9 | .344 |
| Preoperative diameter of the ascending aorta (cm) | 6.0 ± 0.8 | 6.8 ± 0.7 | 5.9 ± 0.8 | .015 |
| Diagnosis of MFS after surgery/all RAA for aneurysm >5.0 cm | 6/36 (16.7) | 0/4 | 6/32 (18.8) | 1.000 |
| Delayed RAA for ATAAD | 42 (14.6) | 0 | 42 (24.9) | <.001 |
| Age at surgery (years) | 37.7 ± 10 |  | 37.7 ± 10 |  |
| Diagnosis of MFS after surgery/all ATAAD repairs | 34/42 |  | 34/42 |  |
| All RAA procedures at or after the age of 21 years | 141 (49) | 25 (21) | 116 (68.6) | <.001 |
| Age at surgery (years) | 3.8 ± 1.1 | 26.7 ± 11.2 | 38.5 ± 11.8 | <.001 |
| Diagnosis of MFS after surgery/all RAA procedures | 42/141 (29.8) | 0/25 | 42/116 (26.2) | <.001 |
| Surgical techniques all RAA procedures |  |  |  | .056 |
| • Supra-coronary replacement | 10 | 0 | 10 |  |
| • Composite grafting (Bentall) | 41 | 4 | 37 |  |
| • Aortic-valve-sparing technique (David) | 90 | 21 | 69 |  |

RAA, replacement of the ascending aorta; ATAAD, acute type A aortic dissection, n = number of patients
\*Kruskal-Wallis test or Fisher's generalized exact test

### Primary outcome

Kaplan-Meier curve analysis showed a higher risk for delayed RAA with late versus early MFS diagnosis. At the age of 50 years, only 9% with early vs 44% with late MFS diagnosis underwent delayed RAA (log-rank P<0.001; Figure 2). Delayed RAA for ATAAD was performed only with late MFS diagnosis. Multivariate analysis identified late MFS diagnosis as an independent predictor of the primary outcome (hazard ratio [HR] 7.39; 95% CI 2.68 to 20.37; P<0.001) and confirmed male sex as an additional predictor (HR 1.72; 95%CI 1.09 to 2.71; P=0.020; Figure 3).

**Figure 2.**
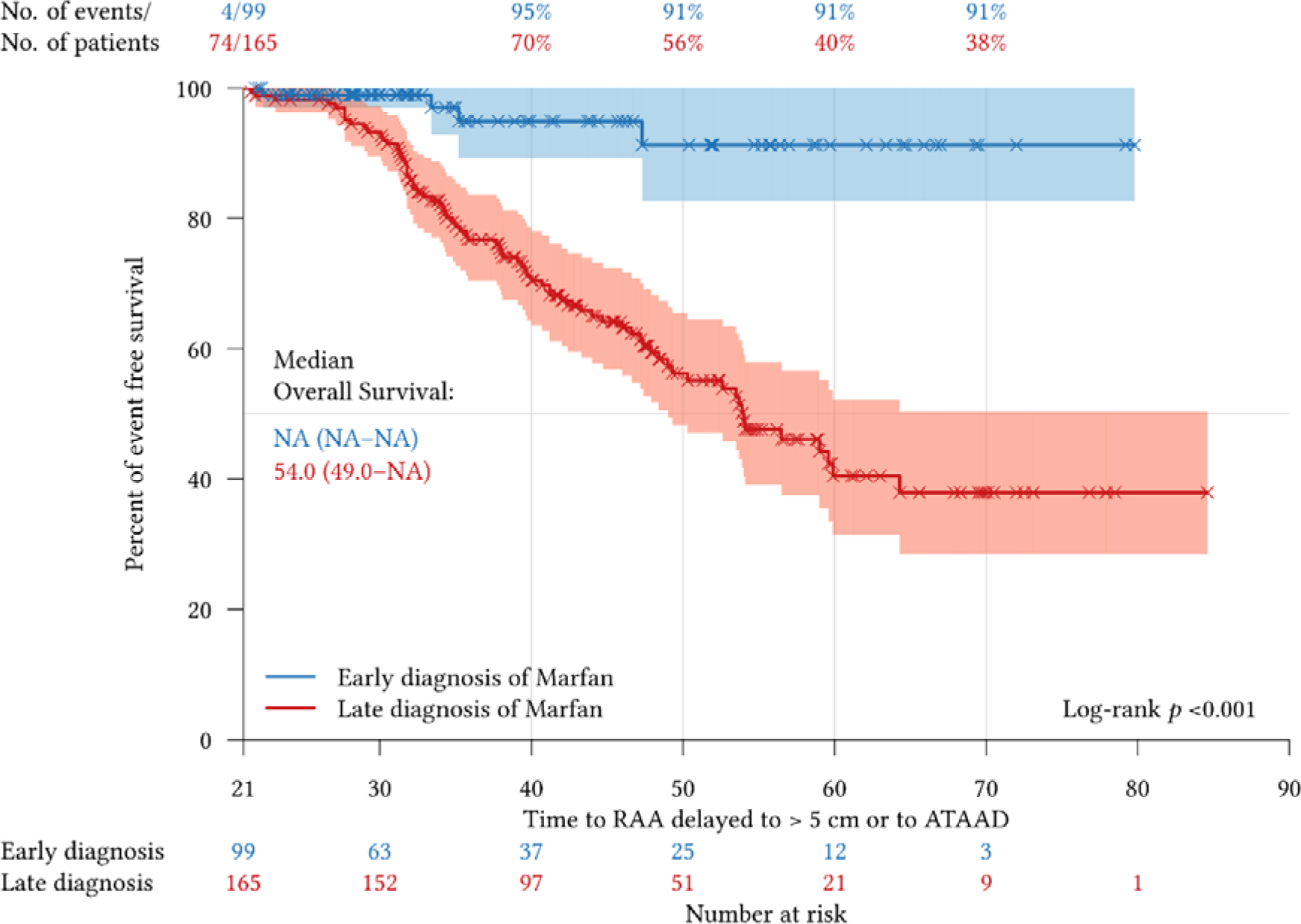
Kaplan-Meier estimates of primary outcome. The time-to event analysis from landmark age 21 years to primary outcome, defined as delayed replacement of the ascending aorta (RAA) for diameters >5.0cm or for acute type A aortic dissection. Comparisons were made for early (age <21 years) versus late diagnosis of Marfan syndrome (age ≥21 years).

**Figure 3.**
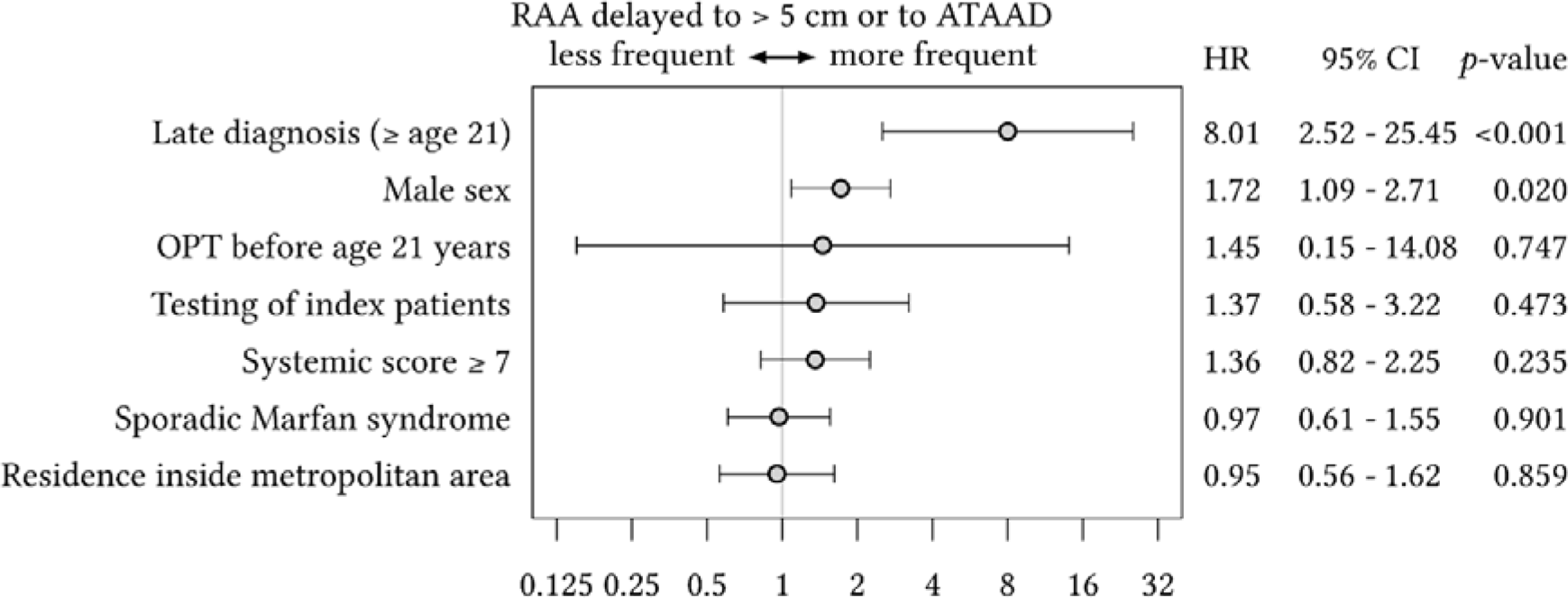
Forest plot of multivariate Cox regression analysis of primary outcome. The primary outcome was delayed replacement of the ascending aorta (RAA) for aortic diameters >5.0cm or for acute type A aortic dissection (ATAAD). Multivariate Cox regression analysis examined the independent association of late diagnosis of Marfan syndrome and late optimal pharmacological therapy (OPT), both defined by age ≥21 years, with primary outcomes after adjustment for 5 other study variables with known risk of aortic events. The grey vertical line represents the hazard ratio for the overall study population.

### Secondary outcome

Kaplan-Meier curve analysis revealed higher risk for death with late versus early MFS diagnosis (log-rank P<0.025; Figure 4). Multivariate analysis revealed late MFS diagnosis as an independent predictor of death (HR 4.68; 95% CI 1.17 to 18.80; P=0.029), and RAA before the age of 21 years as an additional predictor (HR 14.58; 95% CI 3.80 to 55.85; P<0.001; Figure 5). Kaplan-Meier curve analysis illustrates the higher risk of death with RAA before the age of 21 years (log-rank P<0.001; Figure S2).

**Figure 4.**
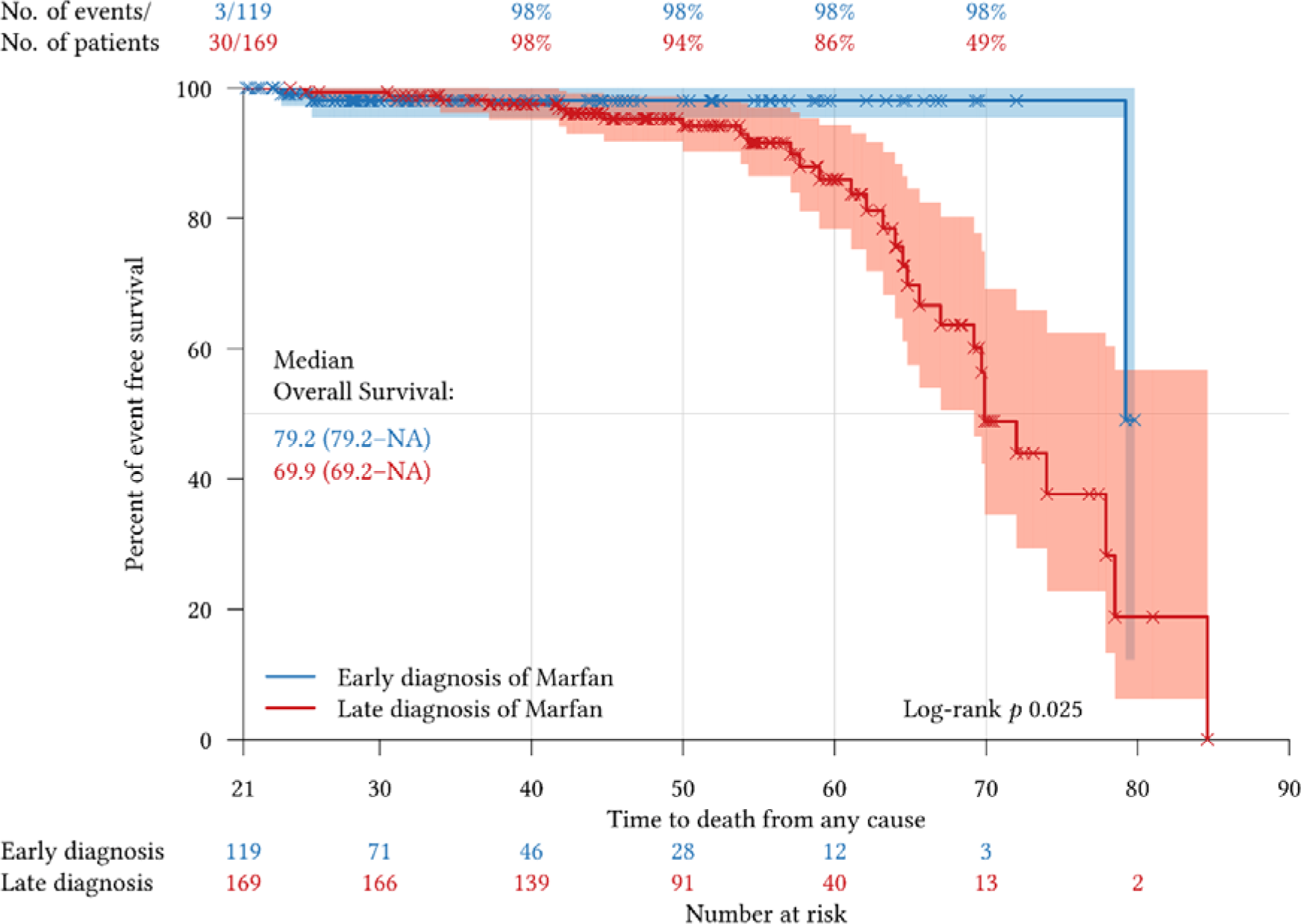
Kaplan-Meier estimates of secondary outcome. The time-to event analysis from landmark age 21 years to secondary outcome, defined as all-cause mortality. Comparisons were made for early (age <21 years) versus late diagnosis of Marfan syndrome (age ≥21 years).

**Figure 5.**
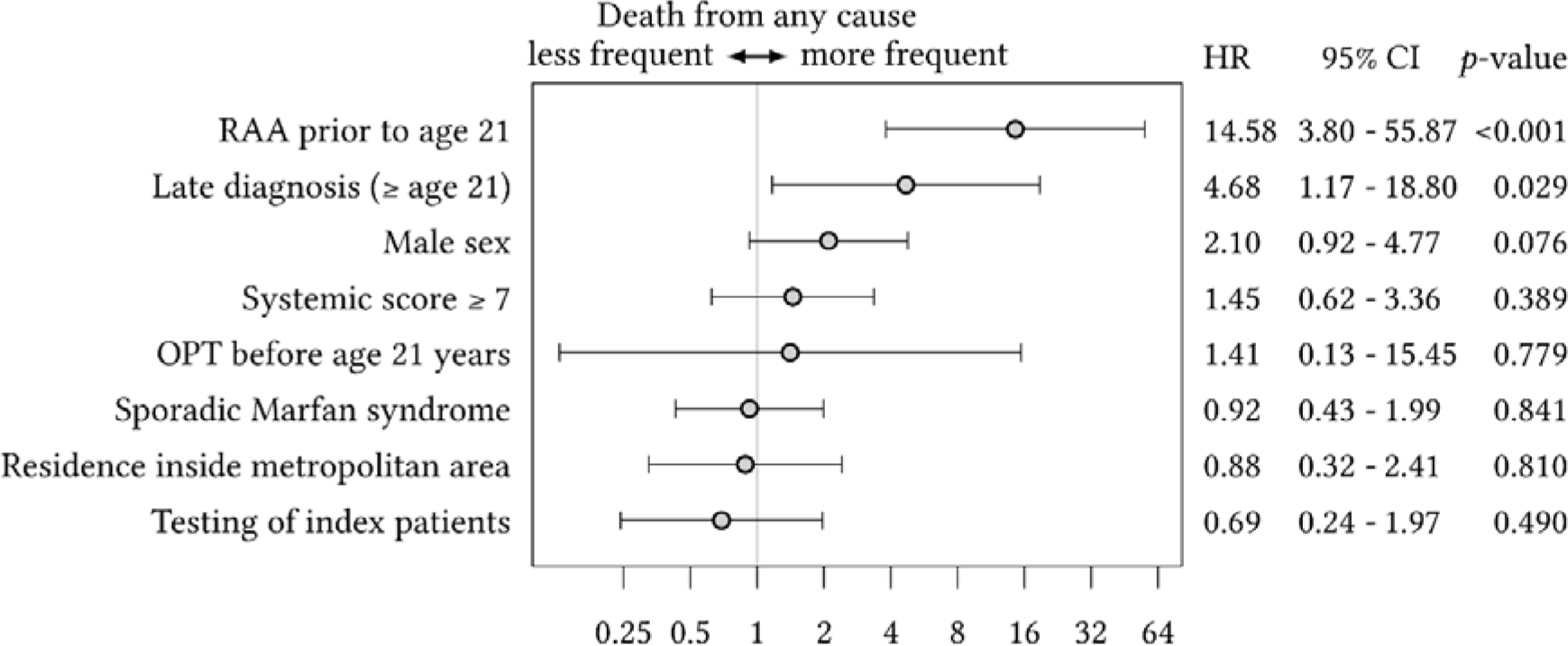
Forest plot of multivariate Cox regression analysis of secondary outcome. The secondary outcome was all-cause mortality. Multivariate Cox regression analysis examined the independent association of late diagnosis of Marfan syndrome and late optimal pharmacological therapy (OPT) initiation, both defined by age ≥21 years, with secondary outcomes after adjustment for 5 other study variables with known risk of aortic events. The grey vertical line represents the hazard ratio for the overall study population, ARR identifies replacement of the ascending aorta.

### Sub-analyses

First, we analyzed the primary outcome separately for RAA for aneurysms >5.0cm and for ATAAD. Kaplan-Meier curve analysis revealed higher risk with late versus early MFS diagnosis for both, delayed RAA for aneurysms >5.0cm (log-rank P=0.040; Figure S3), and for delayed RAA for ATAAD (P<0.001; Figure S4). Multivariate Cox regression analysis confirmed a marginal association of late MFS diagnosis with delayed RAA for aneurysms >5.0cm (HR 2.71; 95% CI 0.94 to 7.81; P=0.065; Figure S5). Cox regression analysis was not performed for delayed RAA for ATAAD, because there were no events with early MFS diagnosis.

Second, we analyzed MFS diagnosis by the time before or after RAA and by the time before death. Diagnosis was established before RAA in all 44 RAA with early MFS diagnosis, whereas in the late MFS diagnosis group, MFS was diagnosed after RAA in all 5 RAA before the age of 21 years, in 2 of 44 timely RAA at diameters ≤5.0cm (2.9%), in 6 of 32 delayed RAA for aneurysms >5.0cm (18.8%), and in 33 of 42 delayed RAA for ATAAD (78.6%; P<0.001; Table 3; Figure S6, upper panel). The 113 survivors with early MFS diagnosis had a significantly longer life with MFS diagnosis (23±15.5 years) than 149 survivors (11.4±15.5 years) and 24 decedents with late MFS diagnosis (10±6.4 years; P<0.001; Figure S6, lower panel).

Third, we analyzed causes of death. All deaths were cardiovascular, with the immediate cause of death being sudden unexplained death in 17 patients, heart failure or aortic disease in 7 patients each, and sepsis after cardiovascular surgery in 2 patients. Analysis of deaths by initial cardiovascular procedure showed that 13.9% of delayed RAA for aneurysms >5.0cm, 17.3% of RAA before age 21 years, and 26.2% of delayed RAA for ATAAD ultimately resulted in death. Before their final death, approximately 50% of these patients required at least one additional cardiovascular procedure (Table S5).

Finally, for validation of outcomes we compared the current study cohort of 341 patients with MFS with data from German health insurance claims. The overall mortality was comparable to that of MFS and it was significantly lower than that of degenerative aortic disease (Figure S7).

## Discussion

To our knowledge, this is the first study to examine the effects of a late MFS diagnosis on long-term outcomes. Our main finding is that a late diagnosis of MFS is associated with higher rates of urgent and emergent aortic operations, and higher cardiovascular mortality. Our findings reinforce recommendations for early detection of MFS. The study cohort and outcomes were representative of MFS in the general population (2). Late diagnosis of MFS, defined as at age ≥21 years based on aortic risk and the median age of MFS diagnosis in a national cohort, was common in our study cohort (41.3%) (3). A late MFS diagnosis related to genetic testing in index patients rather than in family members, and to milder MFS phenotypes (4). Late diagnosis carried an 8.01-fold risk of delayed RAA (5) and a 2.8-fold risk of death (6). These results underscore the importance of research and interventions to reduce late MFS diagnosis (6). The subjects (1-6) are discussed below by number.

### 1. Scoping review

The scoping review of Medline extracted 119 records with statements on the prognostic impact of the time of diagnosis of MFS (Figure S8). In 113 of these reports, the prognostic significance of the time of diagnosis was emphasized, however without direct evidence. Only 6 studies measured the time of diagnosis ^6, 11, 16–19^: One study used questionnaires to examine the time between the first symptoms and the final diagnosis in 430 patients ^16^. Another retrospective study of 66 adults with MFS compared outcomes of 27 patients with a diagnosis before and 39 with a diagnosis after the onset of adulthood ^17^. In a study of administrative data from 389 patients with MFS, the delay in diagnosis was measured in days from the first symptoms to the final diagnosis ^18^. Finally, three Danish studies with administrative data reported age at initial diagnosis ^11^, time of diagnosis in relation to aortic events ^6^, and pregnancy (Table S6) ^19^. EURORDIS, not listed in Medline, measured diagnostic delays using questionnaire responses of 682 families with MFS ^20^. Therefore, this scoping review documents consensus on the prognostic importance of time at diagnosis, but sparse data, heterogeneous methods and varying definitions were inappropriate for time-to-event analyses.

### 2. Representativeness of the cohort and outcomes

The proportion of patients with MFS in the study cohort to the total number of inhabitants in the Hamburg metropolitan area equalled 5.7 patients per 100,000 inhabitants, which was close to the prevalence of 6.5 individuals with MFS per 100,000 in the Danish population ^11^. Comparison of the current study cohort of 341 patients with MFS with data from German health insurance claims revealed an overall mortality comparable to that of MFS and significantly lower than that of degenerative aortic disease (Figure S7).

### 3. Commonness of late diagnosis

In this study, the median age of 20 years at diagnosis was high, but comparable to 19 years median age in the Danish study ^11^. Similarly, the time between first consultation to diagnosis was 2.8 years in the French study ^16^, 2.0 years in 50% of patients in EURORDIS ^20^, and a median of 641 days in the German study ^18^. Therefore, the widespread prevalence of late MFS diagnosis is a well-documented problem.

### 4. Causes of late diagnosis

Genetic cascade testing can diagnose MFS young family members of index patients with MFS despite their own mild phenotype. However, cascade testing was performed less frequently in late (11.8%) than in early MFS diagnosis (31.1%; P<0.001). Moreover, severe MFS phenotypes, as indicated by systemic Ghent score ≥7 points, RAA before the age of 21 years, and younger age at timely RAA (≤5.0cm), were more common in early vs late MFS diagnosis. Therefore, less cascade testing and milder phenotypes related to late MFS diagnosis.

### 5. Causes and consequences of delayed RAA

With a hazard ratio >4.25, the independent effect of late MFS diagnosis on the risk of delayed RAA was huge in this study ^14^. The risk of a late MFS diagnosis was confirmed by separate analysis of delayed RAA for aneurysm >5.0cm and for ATAAD. The quality of expert care was comparable in early and late MFS diagnosis, as suggested by similar frequency of echocardiography, timely RAA, and number of losses to follow-up. However, expert care started earlier, lasted longer, and lead to OPT before age 21 years only with early diagnosis. Similarly, late MFS diagnosis resulted in delayed RAA, since only with late MFS diagnosis MFS was diagnosed after RAA, with the highest frequency in delayed RAA for ATAAD (81%). Finally, guidelines discourage supra-coronary RAA in MFS ^1^. However, this study documents 11 supra-coronary RAA procedures with late vs none with MFS diagnosis.

### 6. Causes of death

With a hazard ratio >4.25, the independent effect of late MFS diagnosis on the risk of death was also huge ^14^. The majority of deaths occurred after delayed RAA (16/33). In addition, in all 4 patients with surgery of the descending aorta or mitral valve surgery as initial cardiovascular procedure with lethal outcome later on, MFS diagnosis was also delayed until after surgery. This study identified RAA before the age 21 years as another independent predictor of death in MFS, probably because of severe MFS phenotype. Both, in delayed RAA and RAA before the age of 21 years subsequent cardiovascular surgeries were frequent and contributed to ultimate death (Table S5).

### 7. Future interventions

Our data suggest that early diagnosis leads to timely operation with good outcomes in MFS. Our analysis could be the foundation of a controlled trial testing population-based screening for MFS, using genetic analysis, established scores such as the Sheikhzadeh Score ^21^ and the Kid-Short Marfan Score ^22^, or imaging of the aortic diameter to improve outcomes in this condition. Medical education programs for frontline caregivers and public campaigns to raise awareness of rare diseases, dissemination of screening tools, databases and decision support systems, improved coordination of diagnostic processes, strengthening of patient advocacy organizations ^20^, and wider use of genetic testing may be appropriate to enable timely diagnosis of MFS in more patients.

### Study limitations

Strengths of the study are the relatively large sample size, the verification of MFS and its genetic cause at a single reference center, the representativeness of the cohort for MFS in the population, the long follow-up period and careful capture of outcomes. A comprehensive analysis of deaths due to ATAAD in our region confirms that we did not miss any patients who died of MFS-related aortic aneurysm. Weaknesses include the combination of prospective and retrospective data collection and the single reference-center patient recruitment. We aimed to mitigate these weaknesses by adding analyses from National Insurance data suggesting that our data set captures a representative sample of patients with MFS. We applied the GRADE steps to assess the certainty of evidence and calculated a GRADE score of 4 (Table S7, Table S8). Prospective external validation of our findings is desirable, e.g. in other large referral centers ^22^. This analysis is largely limited to patients of European ancestry. Outcomes in other ethnicities may differ. The analysis is also limited to *FBN1*-positive MFS. Results in other hereditary thoracic aortic diseases may differ.

### Conclusions

This study demonstrates that a late diagnosis of MFS is associated with a higher rate of delayed aortic surgery, defined as RAA for aortic aneurysms >5.0cm or RAA for ATAAD, and death. Time of diagnosis of MFS should be considered as a criterion for quality of care. Medical education programs, public awareness campaigns, screening tools, rare disease databases, decision support systems, better coordination of diagnostic procedures, strengthening of patient organizations, and wider use of genetic testing may be appropriate measures to reduce the age at diagnosis of MFS.

## Data Availability

There are no external datasets or supplementary materials online at other repositories that pertain to this manuscript

