## Supplementary material for "Late diagnosis of Marfan syndrome is associated with unplanned aortic surgery and cardiovascular death": Supplemetary file

Supplemental Material

Contents

Table S1. STROBE checklist	2

Table S2. PRISMA-ScR checklist	6

Table S3. Exclusion from study because of age < 21 years	13

Table S4. Study patients by discontinuation of study	15

Table S5. Cardiovascular procedures in study patients with death	16

Table S6. PRISMA-ScR results	17

Table S7. GRADE results	21

Table S8. QIPS tool results	26

Figure S1. Study participants	32

Figure S2. Kaplan-Meier estimates of death according to RAA before age 21 years	33

Figure S3. Kaplan-Meier estimates of delayed RAA for aneurysm > 5.0 cm	34

Figure S4. Kaplan-Meier estimates of delayed RAA for ATAAD	35

Figure S5. Forest plot of multivariate Cox regression analysis of RAA delayed > 5.0 cm	36

Figure S6. MFS diagnosis by the time before or after surgery or death	37

Figure S7. Validation of outcomes	38

Figure S8. PRISMA-ScR 2018 flow diagram	39

References	40

### Table S1. STROBE checklist

STROBE statement - checklist of items that should be included in reports of observational studies ^1^

| Manuscript | Item | Recommendation | Section of manuscript |
| --- | --- | --- | --- |
| Title and abstract | 1 | (a) Indicate the study’s design with a commonly used term in the title or the abstract | Title |
|  |  | (b) Provide in the abstract an informative and balanced summary of what was done and what was found | Abstract |
| Introduction |  |  |  |
| Background/rationale | 2 | Explain the scientific background and rationale for the investigation being reported | Introduction |
| Methods |  |  |  |
| Objectives | 3 | State specific objectives, including any pre-specified hypotheses | Introduction |
| Study design | 4 | Present key elements of study design early in the paper | Methods, patients, diagnostic assessment, and follow-up |
| Study design |  |  |  |
| Variables | 7 | Clearly define all outcomes, exposures, predictors, potential confounders, and effect modifiers. Give diagnostic criteria, if applicable | Methods, outcomes |
| Data source/ measurement | 8 | For each variable of interest, give sources of data and details of methods of assessment (measurement). Describe comparability of assessment methods if there is more than one group | Methods, statistical analysis |
| Bias | 9 | Describe any efforts to address potential sources of bias | Methods, Capture and completeness of events, certainty of evidence |
| Study size | 10 | Explain how the study size was arrived at | Methods, statistical analysis |
| Quantitative variables | 11 | Explain how quantitative variables were handled in the analyses. If applicable, describe which groupings were chosen and why | Methods, statistical analysis |
| Statistical methods | 12 | (*a*) Describe all statistical methods, including those used to control for confounding | Methods, statistical analysis |
|  |  | (*b*) Describe any methods used to examine subgroups and interactions | Methods, statistical analysis |
|  |  | (*c*) Explain how missing data were addressed | Methods, statistical analysis |
|  |  | (*d*) *Cohort study*—If applicable, explain how loss to follow-up was addressed  *Case-control study*—If applicable, explain how matching of cases and controls was addressed  *Cross-sectional study*—If applicable, describe analytical methods taking account of sampling strategy | Methods, outcomes |
|  |  | (*e*) Describe any sensitivity analyses | N/A |
| Results |  |  |  |
| Participants | 13 | (a) Report numbers of individuals at each stage of study—eg numbers potentially eligible, examined for eligibility, confirmed eligible, included in the study, completing follow-up, and analyzed | Results, population, outcomes |
|  |  | (b) Give reasons for non-participation at each stage | Results, Figure 1 |
|  |  | (c) Consider use of a flow diagram | Figure 1 |
| Descriptive data | 14 | (a) Give characteristics of study participants (eg demographic, clinical, social) and information on exposures and potential confounders | Results, study groups, Table 1 |
|  |  | (b) Indicate number of participants with missing data for each variable of interest | No missing data |
|  |  | (c) *Cohort study*—Summarize follow-up time (eg, average and total amount) | Results, outcomes, Table 2 |
| Outcome data | 15 | *Cohort study*—Report numbers of outcome events or summary measures over time | Results, outcomes, Table 2; Figures 3, 5 |
|  |  | *Case-control study—*Report numbers in each exposure category, or summary measures of exposure | N/A |
|  |  | *Cross-sectional study—*Report numbers of outcome events or summary measures | N/A |
| Main results | 16 | (a) Give unadjusted estimates and, if applicable, confounder-adjusted estimates and their precision (eg, 95% confidence interval). Make clear which confounders were adjusted for and why they were included | Results, outcomes, composite endpoints, overall mortality |
|  |  | (b) Report category boundaries when continuous variables were categorized | Results, Figures 3-6 |
|  |  | (c) If relevant, consider translating estimates of relative risk into absolute risk for a meaningful time period | Results, outcomes, composite endpoints, overall mortality |
| Other analyses | 17 | Report other analyses done—eg analyses of subgroups and interactions, and sensitivity analyses | Results, sub-analyses |
| Discussion |  |  |  |
| Key results | 18 | Summarize key results with reference to study objectives | Discussion, first para |
| Limitations | 19 | Discuss limitations of the study, taking into account sources of potential bias or imprecision. Discuss both direction and magnitude of any potential bias | Discussion, limitations, Tables S6-7 |
| Interpretation | 20 | Give a cautious overall interpretation of results considering objectives, limitations, multiplicity of analyses, results from similar studies, and other relevant evidence | Discussion, scoping review, commonness of late diagnosis, causes of late diagnosis |
| Generalizability | 21 | Discuss the generalizability (external validity) of the study results | Discussion, study limits, Tables S6-7 |
| Other information |  |  |  |
| Funding | 22 | Give the source of funding and the role of the funders for the present study and, if applicable, for the original study on which the present article is based | No funding for this study |

### Table S2. PRISMA-ScR checklist

PRISMA-ScR checklist for scoping review (2)

| Section/topic | # | Checklist item | Reported in chapter of the manuscript |
| --- | --- | --- | --- |
| Title | | |  |
| Title | 1 | Identify the report as a scoping review: This scoping review of the literature is only a part of the original study. Therefore, we identify this review as scoping review in the methods rather than in the title of this manuscript. | Methods, certainty of evidence |
| Abstract | | |  |
| Structured summary | 2 | Provide a structured summary that includes (as applicable): background, objectives, eligibility criteria, sources of evidence, charting methods, results, and conclusions that relate to the review questions and objectives: We provide a structured summary of this scoping review as part of the discussion of our original study. | Discussion, commonness of late diagnosis |
| Introduction | | |  |
| Rationale | 3 | Describe the rationale for the review in the context of what is already known. Explain why the review questions/objectives lend themselves to a scoping review approach | Methods, certainty of evidence |
|  |  | Background: To date, the association between late diagnosis of Marfan syndrome for delay of aortic root replacement, acute type A aortic dissection and death as not been assessed systematically. |  |
|  |  | Rationale: The time of diagnosis of Marfan syndrome may have an impact on the delay of aortic root replacement, acute type A aortic dissection and death. Therefore, we performed a brief scoping review to qualify the nature of evidence on the prognostic relevance of the time of diagnosis of Marfan syndrome |  |
| Objectives | 4 | Provide an explicit statement of the questions and objectives being addressed with reference to their key elements (e.g., population or participants, concepts, and context) or other relevant key elements used to conceptualize the review questions and/or objectives: | Methods, certainty of evidence |
|  |  | Objectives: To assess the evidence on the impact of the age at diagnosis, or a late diagnosis or a delayed diagnosis of Marfan syndrome on outcomes such as aortic events or death.  Population: We restricted this analysis to patients with a diagnosis of Marfan syndrome according to any of the international nosologies of Marfan syndrome including the Berlin nosology ^2^, the first Ghent nosology ^3^ and the revised Ghent nosology ^4^ |  |
| Methods | | |  |
| Protocol and registration | 5 | Indicate whether a review protocol exists; state if and where it can be accessed (e.g., a Web address); and if available, provide registration information, including the registration number: | Methods, certainty of evidence |
|  |  | We considered all scientific publication with information on the time of diagnosis of Marfan syndrome, and its prognostic relevance. |  |
|  |  | We classified the literature according to the type of publication as review articles, case reports, case series, or cohort studies to qualify the certainty of evidence from the literature for the impact of the time of diagnosis of Marfan syndrome on outcomes. |  |
|  |  | We do not have a registered protocol for this brief scoping review of the literature. |  |
| Eligibility criteria | 6 | Specify characteristics of the sources of evidence used as eligibility criteria (e.g., years considered, language, and publication status), and provide a rationale. | Methods, certainty of evidence |
|  |  | We considered all scientific publications with statements or information on the time of diagnosis of Marfan syndrome, and its prognostic relevance. |  |
|  |  | Report characteristics: We considered publications from 1988 (as publication date of the first international diagnostic nosology of Marfan syndrome ^2^) up to finalization of this literature review by end of October, 2022. We included all reports from Medline analysis irrespective of the original language and publication status. |  |
|  |  | Rationale: We were liberal about the quality of evidence of primary literature sources because it was our objective to characterize the scope and quality of evidence on this topic. |  |
| Information sources | 7 | Describe all information sources in the search (e.g., databases with dates of coverage and contact with authors to identify additional sources), as well as the date the most recent search was executed. | Methods, certainty of evidence |
|  |  | Databases with dates of coverage: Medline, from 01.01.1988 to 08.10.2022. |  |
|  |  | We did not contact authors for additional sources of evidence. |  |
|  |  | The final search of literature was executed on 08.10.2022 |  |
| Search | 8 | Present the full electronic search strategy for at least 1 database, including any limits used, such that it could be repeated. | Methods, certainty of evidence,  Table S5, Figure S5 in this Supplementary Appendix |
|  |  | We did not apply search filters to retrieve only specific types of publications, natural language processing or text frequency analysis tools to identify or refine keywords, synonyms, or subject indexing terms, or peer review processes for our search strategy. |  |
|  |  | Exclusion of reports on other diseases than Marfan syndrome, such as neonatal Marfan syndrome or syndromic variants of Marfan syndrome such as Loeys-Dietz syndrome, Ehlers-Danlos syndrome, or MASS phenotype.  Exclusion of reports on Marfan syndrome that did not address the topic of our scoping review  Exclusion of reports for other reasons such as missing information on essential items of the report including abstract or accurate information on the disease or outcomes. |  |
|  |  | The full electronic search strategy including search terms, number or reports retrieved, number or reports excluded and reasons for their exclusion are provided in results table of this scoping review (see |  |
| Selection of sources of evidence | 9 | State the process for selecting sources of evidence (i.e., screening and eligibility) included in the scoping review. | Methods, certainty of evidence |
|  |  | We ran two searches of the literature. The first search was carried out to establish our final search terms and categories of literature. For the final search we employed EndNote software to identify duplicate findings, and for categorizing and counting of the literature by previously established categories. The final EndNote file can be made available upon request to the corresponding author of this study. |  |
| Data charting process | 10 | Describe the methods of charting data from the included sources of evidence (e.g., calibrated forms or forms that have been tested by the team before their use, and whether data charting was done independently or in duplicate) and any processes for obtaining and confirming data from investigators. | Methods, certainty of evidence |
|  |  | We applied the criteria and classifications of literature as established during the first, preliminary search of the literature for a structured analysis of the literature in the final analysis. We used EndNote groups to document the categorization of each of the total of 601 publications screened for this report. |  |
| Data items | 11 | List and define all variables for which data were sought and any assumptions and simplifications made. | Methods, certainty of evidence, Table S5 in this Supplementary Appendix |
|  |  | We provide the complete list of search terms in the results of our scoping review |  |
| Critical appraisal of individual sources of evidence | 12 | If done, provide a rationale for conducting a critical appraisal of included sources of evidence; describe the methods used and how this information was used in any data synthesis (if appropriate). | N/A |
| Synthesis of results | 13 | Describe the methods of handling and summarizing the data that were charted. | Methods, certainty of evidence, Table S5 in this Supplementary Appendix |
| Results |  |  |  |
| Selection of sources of evidence | 14 | Give numbers of sources of evidence screened, assessed for eligibility, and included in the review, with reasons for exclusions at each stage, ideally using a flow diagram. | Table S5 in this Supplementary Appendix |
| Characteristics of sources of evidence | 15 | For each source of evidence, present characteristics for which data were charted and provide the citations. | Table S5 in this Supplementary Appendix |
| Critical appraisal within sources of evidence | 16 | If done, present data on critical appraisal of included sources of evidence (see item 12): We present the final results of the scoping review as part of the discussion of our study. In this discussion we provide a qualitative estimate of the quality of evidence identified in the literature. | Table S5 in this Supplementary Appendix |
| Results of individual sources of evidence | 17 | For each included source of evidence, present the relevant data that were charted that relate to the review questions and objectives: We summarize the results from review articles, and all kinds of original publications with only indirect evidence only numerically. We present the specific finds from the original literature that actually provides data on the time of diagnosis of Marfan syndrome and its relationship to prognosis in the discussion. | Discussion, commonness of late diagnosis, Table S5 in this Supplementary Appendix |
| Synthesis of results | 18 | Summarize and/or present the charting results as they relate to the review questions and objectives. | Discussion, commonness of late diagnosis, Table S5 in this Supplementary Appendix |
| Discussion |  |  |  |
| Summary of evidence | 19 | Summarize the main results (including an overview of concepts, themes, and types of evidence available), link to the review questions and objectives, and consider the relevance to key groups. | Discussion, commonness of late diagnosis, Table S5 in this Supplementary Appendix |
| Limitations | 20 | Discuss the limitations of the scoping review process. |  |
| Conclusions | 21 | Provide a general interpretation of the results with respect to the review questions and objectives, as well as potential implications and/or next steps. |  |
| Funding |  |  |  |
| Funding | 22 | Describe sources of funding for the included sources of evidence, as well as sources of funding for the scoping review. Describe the role of the funders of the scoping review. | No funding was received for this scoping review of the literature |

### Table S3. Exclusion from study because of age < 21 years

|  |  | Study patients ^a^ | |
| --- | --- | --- | --- |
| Variables | Exclusion for age < 21  (n = 53) | Early diagnosis  (n = 119) | Late diagnosis  (n = 169) |
| Age at diagnosis of Marfan syndrome (years) | 7.3 ± 4.3 | +6.2 | +31.9 |
| Age at first contact with VASERN-GE (years) | 6.3 ± 4.2 | +20.2 | +35.1 |
| Time of care at VASERN-GE (years) | 7.9 ± 2.5 | +4.1 | +2.2 |
| Lost to follow-up | 7 (13.2) | -7.3 | -2.0 |
| Male sex | 29 (54.7) | -13.5 | -6.8 |
| Systemic score ≥ 7 points | 19 (35.8) | +44.9 | +34 |
| OPT before the age of 21 years | 39 (73.6) | -21.5 | -73 |
| RAA before the age of 21 years | 2 (3.8) ^b^ | +12.2 | -1.4 |
| Sporadic occurrence of Marfan syndrome | 17 (32.1) | +9.9 | +10.5 |
| Genetic testing of index patients | 29 (54.7) | +14.2 | +33.5 |
| Residence inside metropolitan area | 39 (73.6) | +0.3 | +2.7 |
| Aortic events at age < 21 years | 0 | +2.6 ^c^ | +1.2 ^d^ |
| RAA, replacement of the ascending aorta; ATAAD, acute type A aortic dissection; n = number of patients, VASCERN-GE, European Reference Network on Rare Multisystemic Vascular Diseases, Germany.  ^a^ Numbers presented as difference in percentage for categorical data and as difference of means for continuous data.  ^b^ One patient with supra-coronary replacement of the ascending aorta, one patient with David operation.  ^c^ One patient with RAA >5.0 cm at age 6-10 years, and two patients ate age 16-20 years of age with aortic root diameters of 5.6, 8.0, and 6.0 cm, respectively.  ^d^ One patient with RAA >5.0 cm at age 11-15 years with an aortic root diameter of 6.0 cm, and one patient with ATAAD at 11-15 years of age. | | | |

### Table S4. Study patients by discontinuation of study

|  | | Discontinuation of study  (study attrition) | |  |
| --- | --- | --- | --- | --- |
| Variables | | Absent  (n = 262) | Present  (n = 26) | P ^a^ |
| Age at initial diagnosis of Marfan-syndrome (years) | | 28.3 ± 15.9 | 31 ± 15.4 | .305 |
| Male sex | | 117 (44.7) | 13 (50) | .681 |
| Systemic score ≥ 7 points | | 196 (74.8) | 18 (69.2) | .638 |
| OPT before the age of 21 years | | 61 (23.3) | 2 (7.7) | .081 |
| RAA before the age of 21 years | | 23 (8.8) | 0 | .244 |
| Sporadic occurrence of Marfan syndrome | | 109 (41.6) | 13 (50) | .414 |
| Genetic testing of index patients | | 205 (78.2) | 26 | .004 |
| Residence inside metropolitan area | | 200 (76.3) | 17 (65.4) | .235 |
| Reasons for not attending regular appointments | |  |  |  |
|  | Moving to a new residence |  | 7 (26.9 ) |  |
|  | Missing appointments |  | 3 (11.5 ) |  |
|  | Not appear before or after cardiovascular surgery |  | 4 (15.4 ) |  |
|  | Receiving care for other severe disease |  | 2 (7.7 ) |  |
|  | Psychiatric disease |  | 1 (3.8) |  |
|  | No specific reasons identified |  | 9 (34.6 ) |  |
| ^a^ Kruskal-Wallis test or Fisher’s generalized exact test | | | | |

### Table S5. Cardiovascular procedures in study patients with death

|  | | Initial procedure: | Subsequent procedures | | | | | |
| --- | --- | --- | --- | --- | --- | --- | --- | --- |
|  | | with post-procedural MFS diagnosis/all  (n = 17/33) | Number | | | Type | | |
| Initial cardiovascular procedure | |  | First  (n = 16) | Second  (n = 7) | Third  (n = 4) | OAR  (n = 16) | TEVAR  (n = 7) | MVR  (n = 4) |
| Procedures at or after age 21 years | |  |  |  |  |  |  |  |
|  | Timely RAA for diameters ≤ 5.0 cm | 2/4 | 2 |  |  |  | 1 | 1 |
|  | Delayed RAA for aneurysm > 5.0 cm | 2/5 | 2 |  |  |  | 2 |  |
|  | Delayed RAA for ATAAD | 9/11 | 6 | 3 | 1 | 8 | 1 | 1* |
|  | OAR of descending or abdominal aorta | 2/2 | 2 | 2 | 1 | 2 | 3 | 0 |
|  | Mitral valve surgery | 2/2 | 0 |  |  |  |  |  |
| RAA before age 21 years | | 0/4 | 4 | 2 | 2 | 6 |  | 2 |
| No cardiovascular procedure | | 0/5† | 0 |  |  |  |  |  |
| RAA, replacement of the ascending aorta; ATAAD, acute type A aortic dissection; MVR, surgical mitral valve repair; n = number of patients; OAR, open aortic repair; TEVAR, thoracic endovascular aortic repair | | | | | | | | |
| *Orthotopic heart transplantation.  †Including one patient with sudden death at 41-45 years of age with aortic root diameter of 5.5 cm before RAA. | | | | | | | | |

### Table S6. PRISMA-ScR results

PRISMA-ScR results of scoping review ^5^

| Medline search terms (01.01.1988 – 08.10.22) | Number of records |
| --- | --- |
| All records according to search | 578 |
| - Marfan syndrome AND diagnostic delay | 88 |
| - Marfan syndrome AND late diagnosis | 123 |
| - Marfan syndrome AND early diagnosis | 361 |
| - Additional records identified through other sources | 6 |
| Records removed because they were duplicate | 82 |
| Records screened by review of the title and abstract of the publication (total) | 496 |
| Records excluded because of the following reasons: | 377 |
| - Other diseases such acute aortic dissection, neonatal Marfan syndrome, other genetic aortic diseases | 146 |
| - Other aspects of Marfan syndrome | 226 |
| - Other reasons | 5 |
| Records with indirect evidence for prognostic role of the time of diagnosis of Marfan syndrome (analysis of title and abstract) | 119 |
| - Review articles that emphasize the importance of an early diagnosis for the prognosis of Marfan syndrome ^6-43^ | 38 |
| - Original work that concludes that an early diagnosis of Marfan syndrome is important for the prognosis ^44-80^ | 37 |
| - Case reports ^81-112^ | 32 |
| - Case series ^113-118^ | 6 |
| Records with direct evidence for prognostic role of the time of diagnosis of Marfan syndrome (analysis of entire article) ^119-124^  Execute summary of all seven records: | 6 |
| Record #1 ^119^: Hirtzlin, I., et al. (2004). "[Medical histories of patients with Marfan's syndrome]." Arch Mal Coeur Vaiss 97(9): 855-860. | |
| The life expectancy of patients with Marfan syndrome has been increased by over 30 years by modern medico-surgical management but the diagnostic problems related to the multiplicity of symptoms and the necessity of collaboration by many specialities complicate the medical history of patients, which is largely unknown. The authors sent a self-administered questionnaire to 1 353 patients with Marfan syndrome to obtain this information. Of the 430 questionnaires returned, as many by men as by women, the diagnosis of the disease was made in less than half the cases by clinical symptoms (42%): the investigation of an unrelated clinical problem (39%) or a family enquiry (19%) also led to the diagnosis. The delay between the first symptoms of the disease and first medical consultation (usually cardiological or ophthalmic) was long (5.2 years) as was the interval between the first consultation and the diagnosis (2.8 years). However, the population could be divided into two groups. one with rapid access to the physician (< 1 year) and to the diagnosis (< 3 years) and the second group in which the delays are long. When the diagnosis of Marfan syndrome is made following consultation for an unrelated condition, it is more often delayed and the patient is usually older. The delay in diagnosis observed could be shortened by systematic familial enquiries and better information to physicians who could then suspect the diagnosis before the advent of clinical symptoms. | |
| Record #2 ^120^: Willis, L., et al. (2009). "Comparison of clinical characteristics and frequency of adverse outcomes in patients with Marfan syndrome diagnosed in adulthood versus childhood." Pediatr Cardiol 30(3): 289-292. | |
| Authors stated that patients with Marfan syndrome (MFS) continue to elude diagnosis until well into adulthood. The purpose of their study was to compare the clinical characteristics and outcomes of adult survivors with MFS diagnosed during adulthood (age, >or=18 years) with those of adult survivors with MFS diagnosed in childhood (<18 years). The investigators conducted a retrospective review of 66 adult (age, >18 years) MFS patients seen at a combined pediatric and adult multidisciplinary MFS clinic from 2004 to 2006. Demographic, clinical, and cardiac variables were collected and a comparative analysis was performed between the two groups: patients diagnosed with MFS during childhood and patients diagnosed in adulthood. The primary outcome measures were the presence of aortic dissection and the need for urgent cardiovascular surgery. Despite a similar incidence of clinical characteristics, 39 of the 66 MFS patients were not diagnosed until adulthood. The overall incidence of major cardiac involvement was comparable between the two groups, although the patients diagnosed at a younger age were found to have a reduced need for aortic surgery (33% vs. 59%; P < 0.04) and fewer adverse cardiac outcomes (0% vs. 46%; P < 0.001). Moreover, the patients diagnosed with MFS in adulthood were more likely to require repeated surgical intervention for distal aortic disease (13% vs. 0%; P = 0.07). In conclusion, patients with MFS who remain undiagnosed until adulthood have well-established cardiovascular pathology frequently requiring surgical intervention. Due to this delay in diagnosis and management, they often suffer from a suboptimal clinical outcome. Our research demonstrates the importance of educating pediatric clinicians in early MFS diagnosis in hopes of improving the long-term outcome of all MFS patients. | |
| Record #3 ^121^: Roll, K. (2012). "The influence of regional health care structures on delay in diagnosis of rare diseases: the case of Marfan Syndrome." Health Policy 105(2-3): 119-127. | |
| This study investigates the relative influence of the regional availability of health care resources (measured by physician densities, number of health care centers) on health care quality (measured by delay in diagnosis), based on data for the rare disease Marfan Syndrome. METHODS: Administrative data from 389 patients with Marfan Syndrome were analyzed. Logistic regression models were applied for a dichotomous comparison of the dependent variable 'time to diagnosis' with the classifications 'immediate' and 'late' diagnosis. Physician densities of cardiologists/angiologists, ophthalmologists, orthopedists, and GPs, as well as distance to medical health care centers and sociodemographic information were entered into the models. RESULTS: The results showed that the relationship between physician densities and probability of immediate diagnosis of Marfan Syndrome is negative linear, and quadratic for cardiologists/angiologists. This effect was significant with respect to density of cardiologists/angiologists (p=0.0097). Distance to medical health care centers was not a predictor of an immediate diagnosis. CONCLUSION: Marfan Syndrome faces significant problems of quality of health care, as although the requisite quantity of health care resources is available, this does not affect delay in diagnosis. Information technology might foster valuable networking among physicians treating such cases along with holistic assessment of symptoms as they occur. | |
| Record #4 ^122^: Groth, K. A., et al. (2015). "Prevalence, incidence, and age at diagnosis in Marfan Syndrome." Orphanet J Rare Dis 10: 153. | |
| The aim of the authors was to study prevalence, incidence, and age at diagnosis in patients with Marfan syndrome. METHOD: Using unique Danish patient-registries, we identified all possible Marfan syndrome patients recorded by the Danish healthcare system (1977-2014). Following, we confirmed or rejected the diagnosis according to the 2010 revised Ghent nosology. RESULTS: The study identified a total of 1628 persons with possible Marfan syndrome. The diagnosis was confirmed in 412, whereof 46 were deceased, yielding a maximum prevalence of 6.5/100,000 at the end of 2014. The annual median incidence was 0.19/100,000 (range: 0.0-0.7) which increased significantly with an incidence rate ratio of 1.03 (95% CI: 1.02-1.04, p < 0.001). The study identified a median age at diagnose of 19.0 years (range: 0.0-74). The age at diagnosis increased during the study period, uninfluenced by the changes in diagnostic criteria. We found no gender differences. CONCLUSION: The increasing prevalence of Marfan syndrome during the study period is possibly due to build-up of a registry. Since early diagnosis is essential in preventing aortic events, diagnosing Marfan syndrome remains a task for both pediatricians and physicians caring for adults. | |
| Record #5 ^123^: Groth, K. A., et al. (2017). "Aortic events in a nationwide Marfan syndrome cohort." Clinical Research in Cardiology 106(2): 105-112. | |
| This study presents aortic event data from a nationwide Marfan syndrome cohort. From the total cohort of 412 patients, 150 (36.4 %) had an aortic event. Fifty percent were event free at age 49.6. Eighty patients (53.3 %) had prophylactic surgery and seventy patients (46.7 %) a dissection. The yearly event rate was 0.02 events/year/patient in the period 1994–2014. Male patients had a significant higher risk of an aortic event at a younger age with a hazard ratio of 1.75 (CI 1.26–2.42, p = 0.001) compared with women. Fifty-three patients (12.9 %) were diagnosed with MFS after their first aortic event which primarily was aortic dissection [n = 44 (83.0 %)]. Conclusion of the authors: More than a third of MFS patients experienced an aortic event and male patients had significantly more aortic events than females. More than half of the total number of dissections was in patients undiagnosed with MFS at the time of their event. This emphasizes that diagnosing MFS is lifesaving and improves mortality risk by reducing the risk of aorta dissection. | |
| Record #6 ^124^: Groth, K. A., et al. (2021). "Maternal health and pregnancy outcome in diagnosed and undiagnosed Marfan syndrome: A registry-based study." Am J Med Genet A 185(5): 1414-1420. | |
| The objective of this study was to demonstrate the consequences on maternal health, in women with diagnosed and undiagnosed MFS at the time of pregnancy and childbirth. By using national health care registries, the study identified all pregnancy related outcomes, from women with MFS (n = 183) and an age-matched background population (n = 18,300). The study found 91 pregnancies during follow-up. Significantly fewer women with MFS gave birth, compared to the background population. No women with known MFS had a pregnancy related aortic dissection but complications related to the cervix were increased (HR:19.8 [95% CI:2.2-177.5]). Fifty women with MFS were undiagnosed at the time of their first pregnancy and/or childbirth. Among these, there were more birth canal related complications HR:27.2 (95% CI: 2.3-315.0), preeclampsia (HR:2.25 [95% CI: 1.11-4.60]), fetal deaths (HR:12.3 [95% CI: 1.51-99.8]), and all delivery-related dissections came from this subgroup. In conclusion, undiagnosed women with MFS experienced more pregnancy and childbirth related complications including fetal death, birth canal issues, preeclampsia, and aortic disease, which emphasizes the need for an early MFS diagnosis and special care during pregnancy and childbirth. | |

### Table S7. GRADE results

GRADE results of grading the evidence for prognostic studies ^125^

|  | Composite endpoint  (RAA > 5.0 cm or ATAAD) | Secondary endpoint  (death of any cause) | GRADE-tool ratings  (for both study endpoints ^126^) |
| --- | --- | --- | --- |
| (1) Phase of investigation ^125^  (instead of study design) | High quality: phase 2 study that provides evidence for the independent association of late diagnosis as prognostic factor with composite study endpoint. | High quality: phase 2 study that provides evidence for the independent association of late diagnosis as prognostic factor with death of all causes. | High quality of evidence counted as four points |
| Factors that can reduce the quality of the evidence ^125^ |  |  |  |
| (2) Limitations in study design or execution (risk of bias according to QIPS tool ^127^): | No serious limitations: Most evidence from this study has a low risk of bias in all bias domains | | No downgrade of the quality of evidence (see Table S7 in this Supplementary Appendix) |
| (3) Inconsistency of results across studies | Inconsistency of study results with the existing body of literature was low, as documented by the scoping review of the literature (see Table S5 in this supplementary appendix). | Not applicable because within the existing body of literature only the present study has estimated the effect of a late diagnosis of MFS as prognostic factor of death. However, it is recommend to downgrade the quality of evidence since this is an indicator that the literature is not well established in the area ^125^. | Downgrade of the quality of evidence by one point |
| (4) Indirectness of evidence | We consider the level of indirectness of evidence as low in all three dimensions of (1) population, (2) prognostic factor, and (3) outcomes: | We consider the level of indirectness of evidence as low in all three dimensions of (1) population, (2) prognostic factor, and (3) outcomes: | No downgrade of the quality of evidence |
|  | (1) Low indirectness of population: The number of patients with MFS was very close to the number of patients expected in the general population. | (1) Low indirectness of population: The number of patients with MFS was very close to the number of patients expected in the general population. | Results, certainty of evidence |
|  | (2) No indirectness in prognostic factor “late diagnosis of MFS”: We applied our definition of “late diagnosis” at diagnosis of MFS ≥ 21 years of age. Of course, other age thresholds could be used, but we report age at diagnosis in all study patients. Age at diagnosis was confirmed in a population-based study in the literature ^122^ | (2) No indirectness in prognostic factor “late diagnosis of MFS”: We applied our definition of “late diagnosis” at diagnosis of MFS ≥ 21 years of age. Of course, other age thresholds could be used, but we report age at diagnosis in all study patients. Age at diagnosis was confirmed in a population-based study in the literature ^122^ | Results, certainty of evidence |
|  | (3) No indirectness of the outcome “composite study endpoint”: We were interested in exploring the prognostic effect of late diagnosis on potentially lethal aortic events of the composite endpoint of surgical delay of RAA to aortic diameters > 5.0 cm or ATAAD. | (3) No indirectness of the outcome “death of all causes”: Death is and remains the most definitive and the most unequivocal endpoint of prognostic studies in diseases with potentially lethal complications such as MFS. | Results, certainty of evidence |
| (5) Imprecision | (1) Sufficient sample size: Sample size was accurately determined (see methods, data analysis) and the desired study power was reached (see results, participants). | (1) Sufficient sample size: Sample size was accurately determined (see methods, data analysis) and the desired study power was reached (see results, participants). | No downgrade of the quality of evidence |
|  | (2) Moderate precision of study: The adjusted 95% confidence interval was 2.52 – 25.45 for the risk of late diagnosis for composite endpoint. | (2) Moderate precision of study: The adjusted 95% confidence interval was 1.17 – 18.80 for the risk of late diagnosis for composite endpoint. |  |
| (6) Publication bias | The scoping review of the literature documents that there are six studies to support the impact of the time of diagnosis on outcomes in Marfan syndrome | The scoping review of the literature documents that there are six studies to support the impact of the time of diagnosis on outcomes in Marfan syndrome | No downgrade of the quality of evidence |
| Factors that can increase the quality of the evidence |  |  |  |
| (1) Large magnitude of effect | The magnitude of the effect of a late diagnosis on composite endpoint was large, defined as hazard ratio > 4.25 ^125^: Adjusted hazard ratio of effect size = 8.01. With 95% confidence interval lying entirely above the null value marked by hazard ratio = 1, this effect also was significant. | The magnitude of the effect of a late diagnosis on death was large, defined as hazard ratio > 4.25 ^125^: Adjusted hazard ratio of effect size = 4.68. With 95% confidence interval lying entirely above the null value marked by hazard ratio = 1, this effect also was significant. | Upgrade of the quality of evidence by one point |
| (2) Exposure-response gradient | Still to be calculated: more years delay of diagnosis since 21 years of age, more risk for composite endpoint? | Still to be calculated: more years delay of diagnosis since 21 years of age, more risk for death? |  |
| Phase-2 studies measure the strength of the prognostic relationship between a factor and the outcome while controlling for alternative explanations ^125^  RAA = replacement of the ascending aorta  ATAAD = acute type A aortic dissection  MFS = Marfan syndrome | | | |
| Comment: We followed all steps (1-8) of the GRADE protocol to estimate the certainty of evidence for the relation of late MFS diagnosis and outcomes. As a first step, this study received 4 points for a high level of evidence because it was a phase 2 explanatory study that established an independent association between late diagnosis and outcomes (1). In a second step, evidence could be downgraded for bias (2), inconsistency (3), indirectness (4), imprecision (5), and publication bias (6). The QUIPS tool excluded study bias, and calculation of population-based prevalence of MFS in our cohort, screening of autopsies for MFS, literature-confirmed incidence of aortic events ^123^, and validation of study mortality with external insurance data all excluded indirect evidence, and the scoping review supported the prognostic significance of timing of diagnosis of MFS (Table S7). Downgrading by one point for death as a study endpoint was necessary because the scoping review did not identify other studies on this topic. In a final step, the evidence was upgraded by 2 points for large hazard ratios >4.25 (7) and exposure-response gradients (8) for each study endpoint. Overall, GRADE scores were ≥4 points with high confidence of evidence for late diagnosis of MFS predicting end points. | | | |

### Table S8. QIPS tool results

QIPS tool results on bias in studies of prognostic factors ^127^

| Criteria | Ratings | Section in the manuscript |
| --- | --- | --- |
| A. Study participation |  |  |
| 1. Source of population of interest | The population of interest is adequately described for all study variables without missing data in any of these variables. Patients excluded for not reaching age ≥ 21 years were analyzed in a distinct sub-analysis. | Results: section participants, Table 1; Table S3 |
| 2. Method used to identify population | The sampling frame and recruitment are adequately described, including methods to identify the sample sufficient to limit potential bias, such as referral patterns. | Methods: patients, diagnostic assessment, and follow-up |
| 3. Recruitment period | Period of recruitment is adequately described | Methods: patients, diagnostic assessment, and follow-up |
| 4. Place of recruitment | Place of recruitment (setting and geographic location) are adequately described | Methods: patients, diagnostic assessment, and follow-up |
| 5. Inclusion and exclusion criteria | Inclusion and exclusion criteria are adequately described (e.g., including explicit diagnostic criteria or “zero time” description). | Methods: patients, diagnostic assessment, and follow-up, Figure 1 |
| 6. Adequate study participation | There is adequate participation in the study by eligible individuals (study power) | Methods: statistical analysis and Results: population |
| 7. Baseline characteristics | The baseline study sample (i.e., individuals entering the study) is adequately described for key characteristics | Results: population, Table 1 |
| Summary study participation | The study sample represents the population of interest on key characteristics, sufficient to limit potential bias of the observed relationship between prognostic factor and outcome. | |
| B. Study Attrition |  |  |
| 1. Proportion of baseline sample available for analysis | Response rate as the proportion of study sample completing the study and providing outcome data is adequate. | Results: section participants; Table 1, Figure 1, failure to attend regular appointments |
| 2. Attempts to collect information on participants who dropped out | Attempts to collect information on participants who dropped out of the study are described. | Methods: outcomes, Table S4 in this Supplementary Appendix |
| 3. Reasons and potential impact of subjects lost to follow-up | Reasons for loss to follow-up are provided. | Results: outcomes;  Table S4 in this Supplementary Appendix |
| 4. Outcome and prognostic factor information on those lost to follow-up | Participants lost to follow-up are adequately described for key characteristics. There are no important differences between key characteristics and outcomes in participants who completed the study and those who did not. | Results: outcomes;  Table S4 in this Supplementary Appendix |
| Study attrition summary | Loss to follow-up is not associated with key characteristics to sufficiently limit potential bias in the observed relationship between prognostic factor and outcome. | |
| C. Prognostic factor measurement |  |  |
| 1. Definition of the prognostic factor | A clear definition or description of 'prognostic factor' is provided by the definition of exposure and non-exposure group of the study | Methods: statistical analysis |
| 2. Valid and reliable measurement of prognostic factor | Method of prognostic factor measurement (age at diagnosis) is adequately valid and reliable to limit misclassification bias. | Methods: statistical analysis |
|  | Continuous variables (age) are reported, and appropriate cut-points for late versus early diagnosis of MFS are used (≥ 21 years of age). | Methods: statistical analysis;  Rationale for age cutoff: see Introduction |
| 3. Method and setting of prognostic factor measurement | The method and setting of measurement of prognostic factor is the same for all study participants. | Methods: Patients, diagnostic assessment, and follow-up |
| 4. Proportion of data on prognostic factor available for analysis | Adequate proportion of the study sample has complete data for prognostic factor variable (no missing data on prognostic variable). | Results: study group;  Table 1 |
| 5. Method used for missing data | Appropriate methods of imputation are used for missing 'prognostic factor' data: No missing data on prognostic factor measurements | Results: study group;  Table 1 |
| Prognostic factor measurement summary | Prognostic factor is adequately measured in study participants to sufficiently limit potential bias | |
| D. Outcome measurement |  |  |
| 1. Definition of the outcome | A clear definition of composite endpoint and all-cause mortality as outcome is provided, including duration of follow-up and level and extent of the outcome construct. | Methods: outcomes; Results: Table 2 |
| 2. Valid and reliable measurement of outcome | The method of outcome measurement for composite endpoint and all-cause mortality is adequately valid and reliable to limit misclassification bias. | Methods: outcomes |
| 3. Method and setting of outcome measurement | The method and setting of outcome measurement is the same for all study participants. | Methods: Patients, diagnostic assessment, and follow-up |
| Outcome measurement summary | Outcome of interest (composite endpoint and all-cause mortality) is adequately measured in study participants to sufficiently limit potential bias. | |
| E. Study confounding |  |  |
| 1. Important confounders measured | Important confounders (study variables as co-variables) and surgical procedures (Table 3) are assessed. | Results: composite endpoints, including Figure 4;  Results: secondary endpoint, including Figure 6 |
| 2. Definition of the confounding factor | Clear definitions of the important confounders measured are provided. | Methods: Statistical analysis |
| 3. Valid and reliable measurement of confounders | Measurement of all important confounders including age, sex, systemic score points, date and technique of surgery, sporadic occurrence of MFS, genetic testing, and place of residence were all measurable in a valid and reliable way. | Methods: Statistical analysis |
| 4. Method and setting of confounding measurement | The method and setting of confounding measurement are the same for all study participants. | Methods: Capture and completeness of events, certainty of evidence |
| 5. Method used for missing data | Appropriate methods are used if imputation is used for missing confounder data, simply because there were no missing data. | Methods: statistical analysis |
| 6. Appropriate accounting for confounding | Important potential confounders are accounted for in the study design (e.g., matching for key variables, stratification, or initial assembly of comparable groups).  Important potential confounders are accounted for in the analysis (i.e., appropriate adjustment). | Methods: Capture and completeness of events, certainty of evidence;  Results: composite endpoints, including Figure 4;  Results: overall survival, including Figure 6 |
| Study confounding summary | Important potential confounders are appropriately accounted for, limiting potential bias with respect to the relationship between prognostic factor and outcome. | |
| F. Statistical Analysis and Reporting |  |  |
| 1. Presentation of analytical strategy | There is sufficient presentation of data to assess the adequacy of the analysis. | Results; including Tables 1-3. |
| 2. Model development strategy | The strategy for model building (i.e., inclusion of variables in the statistical model) is appropriate and is based on a conceptual framework or model. The selected statistical model is adequate for the design of the study. | Methods: statistical analysis |
| 3. Reporting of results | There is no selective reporting of results. | Methods: statistical analysis |
| Statistical analysis and presentation summary | The statistical analysis is appropriate for the design of the study, limiting potential for presentation of invalid or spurious results. | |

### Figure S1. Study participants

STrengthening the Reporting of OBservational studies in Epidemiology (STROBE) flow diagram of the study cohort ^1^.

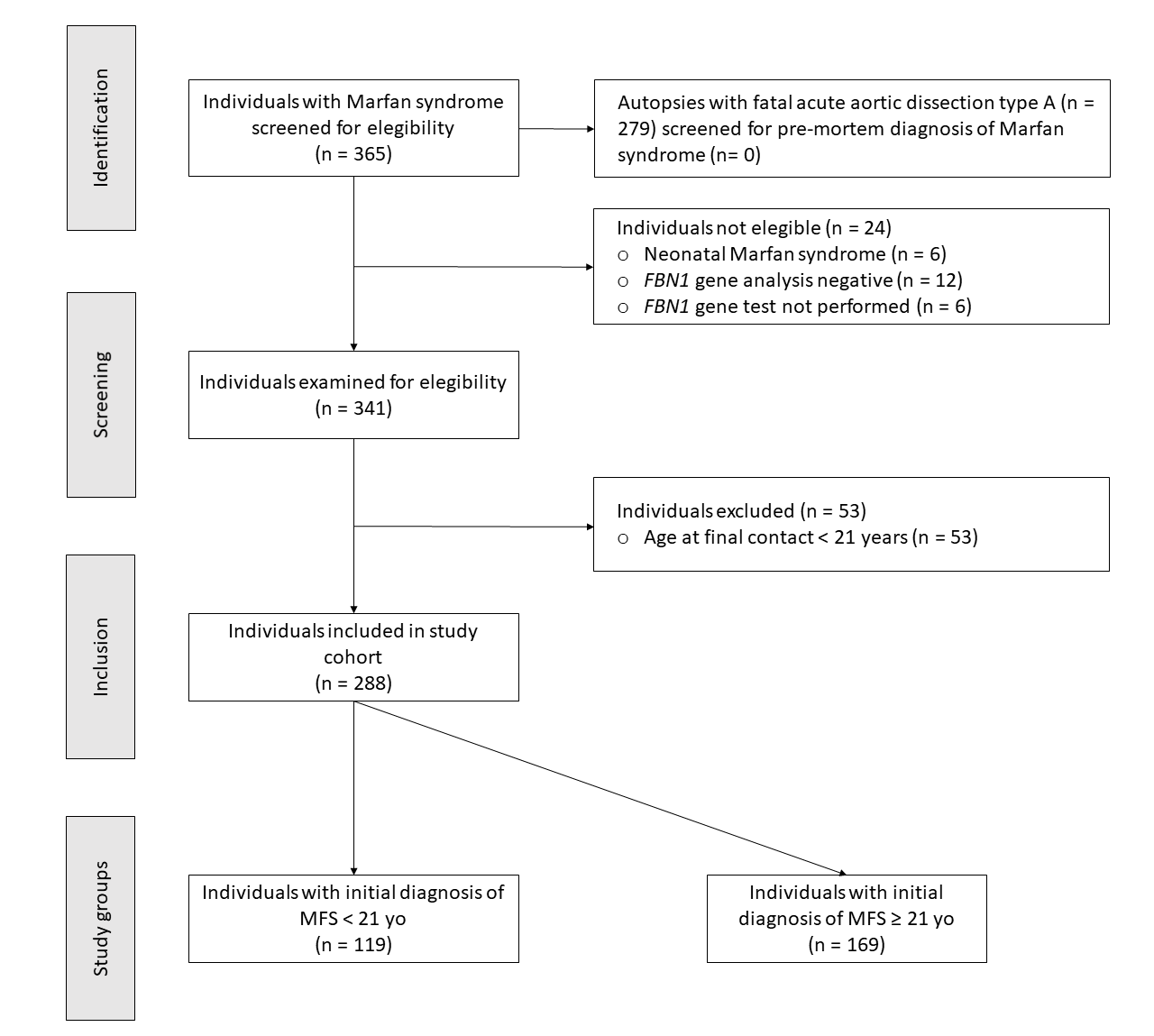

### Figure S2. Kaplan-Meier estimates of death according to RAA before age 21 years

The time-to event analysis from landmark age 21 years to secondary outcome, defined all-cause mortality. Comparisons were made for presence versus absence of RAA before the age 21years.

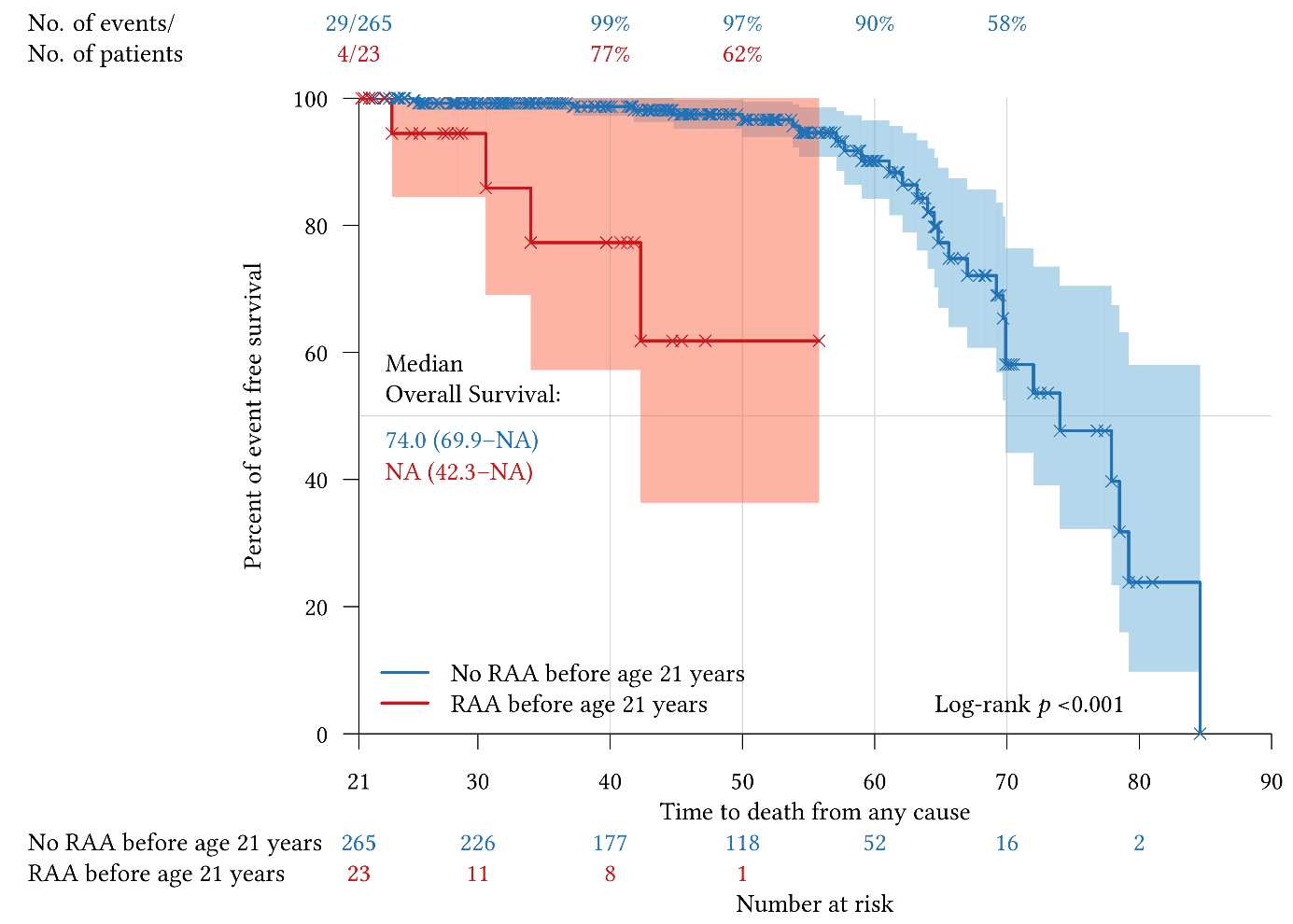

### Figure S3. Kaplan-Meier estimates of delayed RAA for aneurysm > 5.0 cm

The time-to event analysis from landmark age 21 years to delayed aortic root replacement (RAA) aneurysm > 5.0 cm. Comparisons were made for early (age <21 years) versus late diagnosis of Marfan syndrome (age ≥21 years).

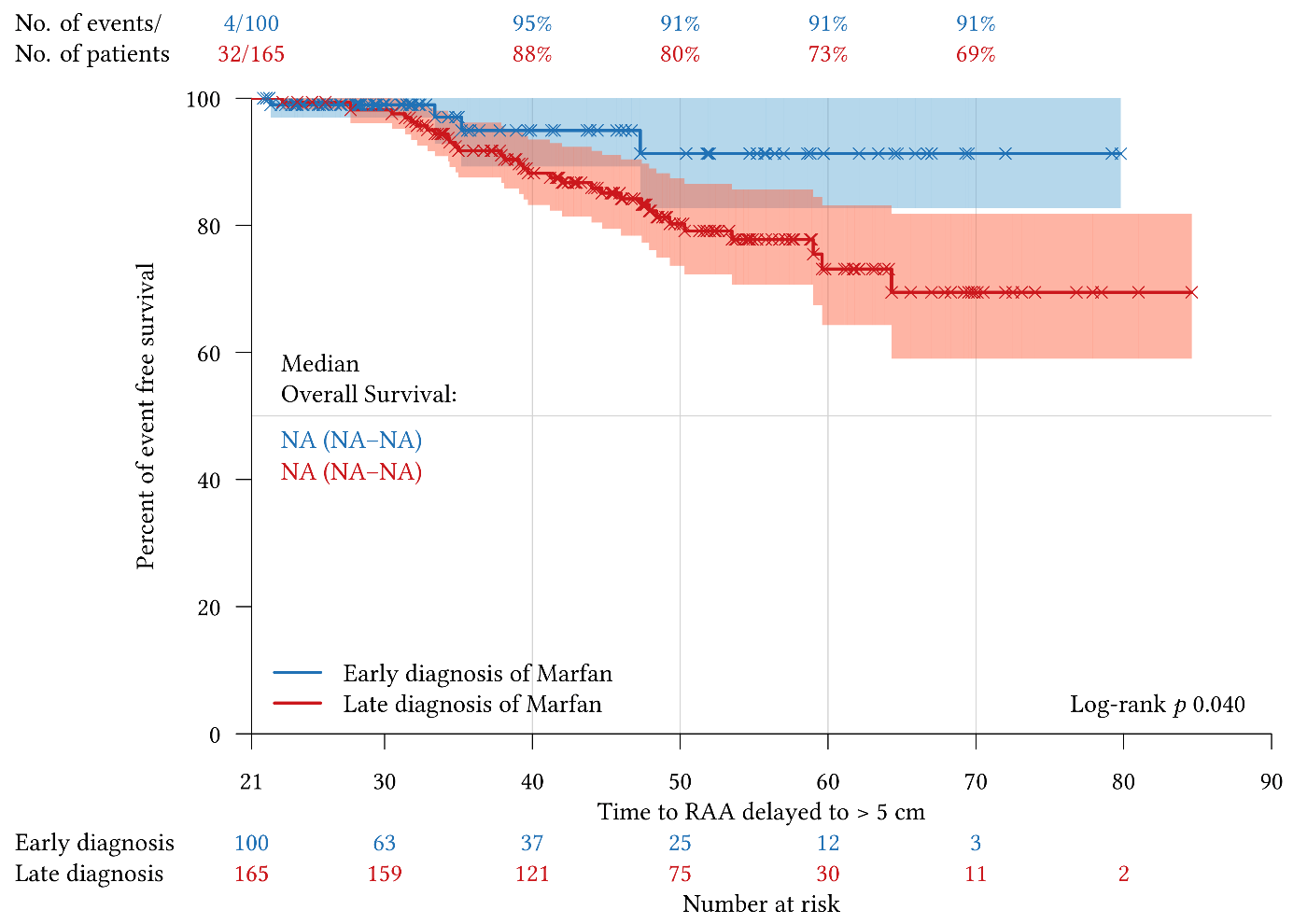

### Figure S4. Kaplan-Meier estimates of delayed RAA for ATAAD

The time-to event analysis from landmark age 21 years to delayed RAA for acute type A aortic dissection (ATAAD). Comparisons were made for early (age <21 years) versus late diagnosis of Marfan syndrome (age ≥21 years).

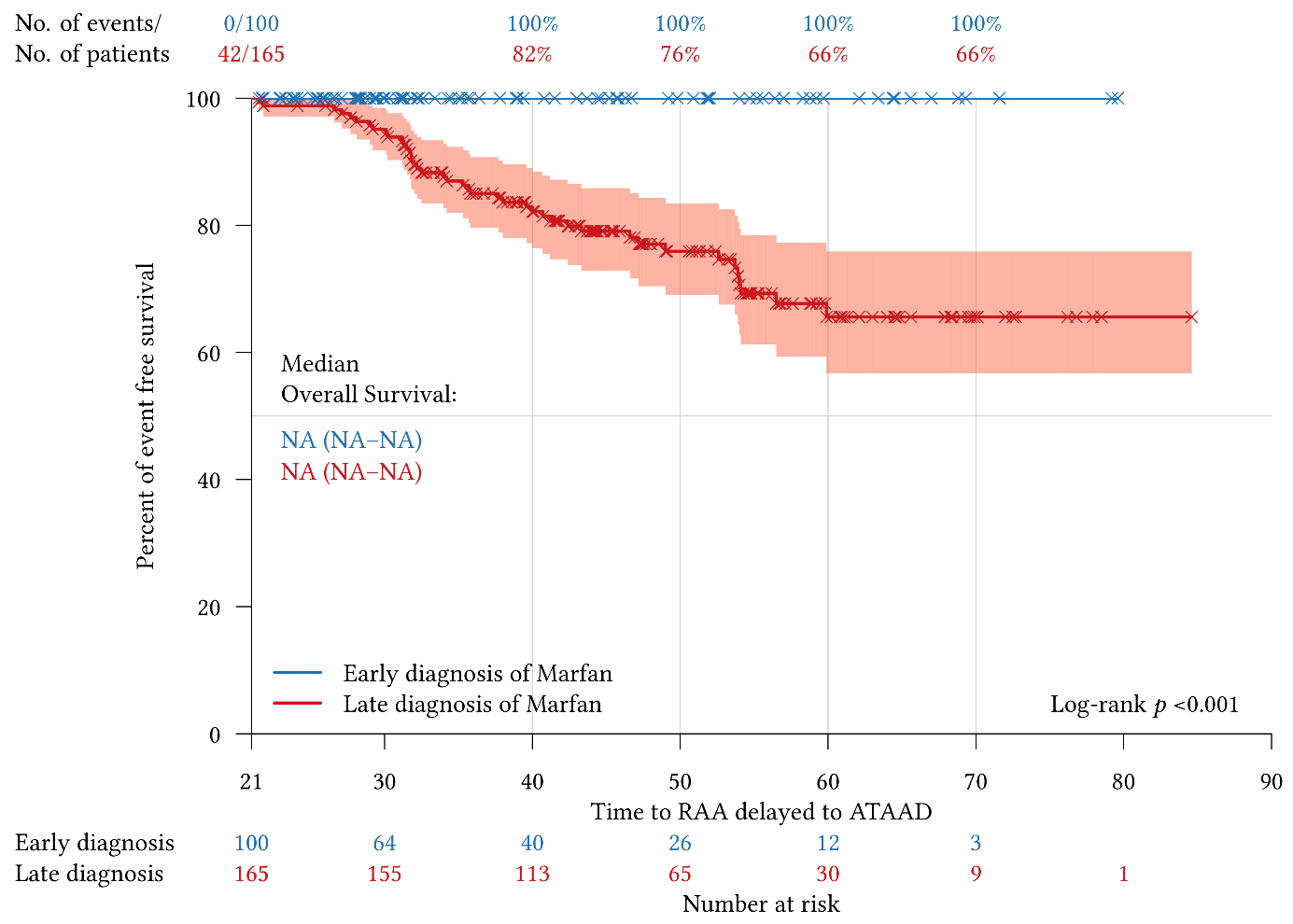

### Figure S5. Forest plot of multivariate Cox regression analysis of RAA delayed > 5.0 cm

The endpoint was delay of aortic root replacement (RAA) to aneurysm > 5.0 cm. Multivariate Cox regression analysis examined the independent association of late diagnosis of Marfan syndrome, defined diagnosis at an age ≥ 21 years, with delayed RAA for aneurysm > 5.0 cm after adjustment for 6 other study variables with known risk of aortic events. The grey vertical line represents the hazard ratio for the overall study population.

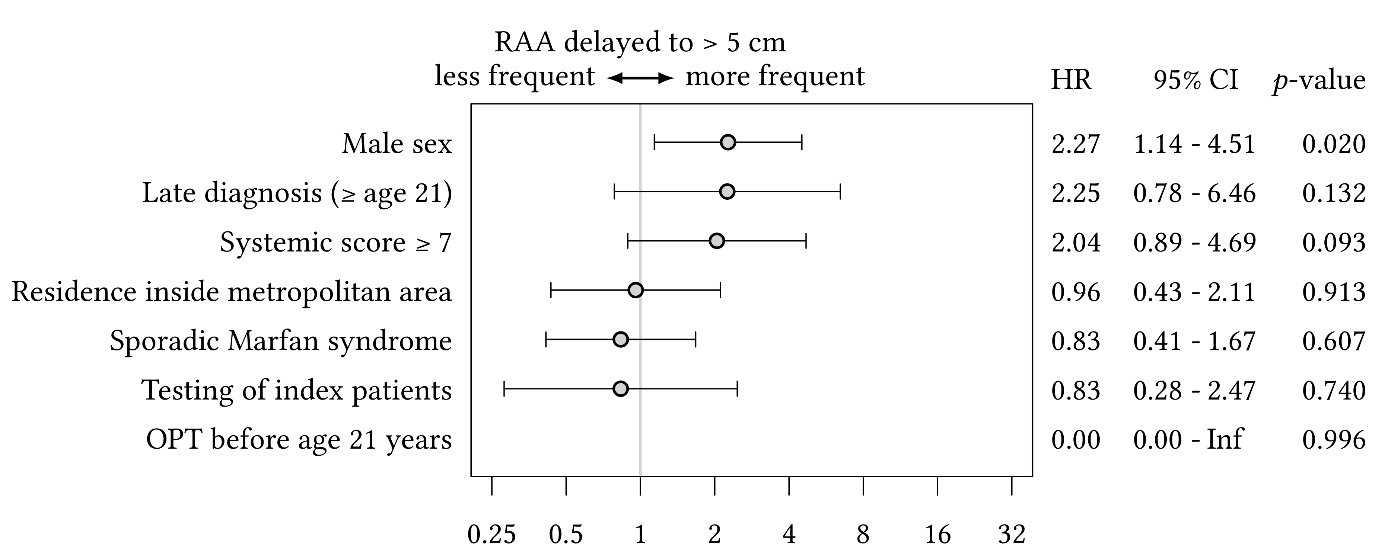

### Figure S6. MFS diagnosis by the time before or after surgery or death

The box and whiskers plots of the time of diagnosis of Marfan syndrome (MFS) before (negative time values) or after replacement of the ascending aorta (positive time values; RAA) in years, according to early versus late diagnosis of Marfan syndrome and the type of surgical procedure (upper panel). RAA ≤5.0cm and >5.0cm denotes aortic root replacement at respective preoperative aortic diameters, and ATAAD acute type A aortic dissection. The same analysis is shown for time of MFS diagnosis in years (negative values) before death (lower panel).

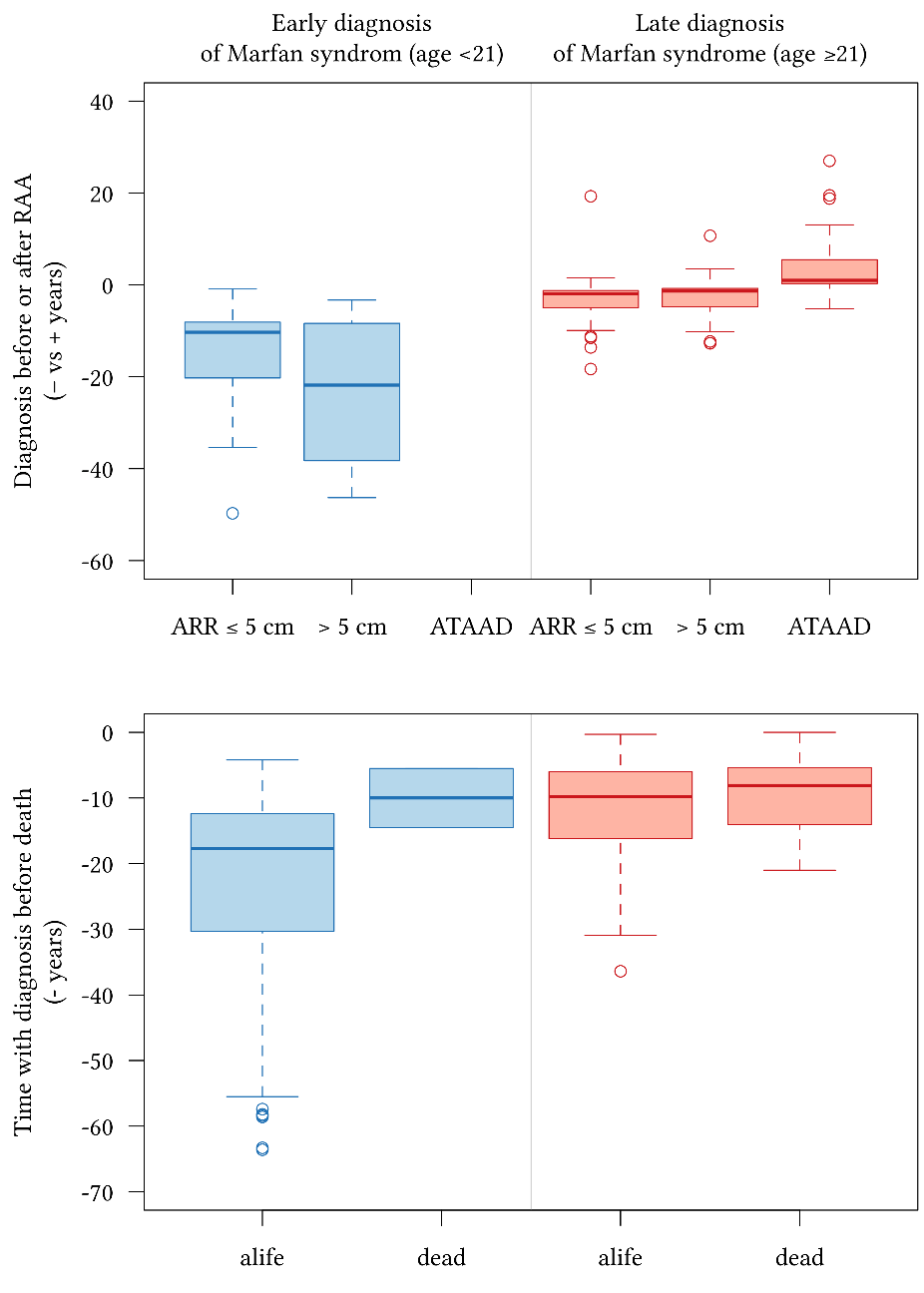

### Figure S7. Validation of outcomes

The outcome of time to event analysis was death of any cause. Patients with Marfan syndrome from the current study cohort were in group C, patients from the DAK-Gesundheit health insurance with ICD Q87.4 coding for MFS were in group B, and patients from the DAK-Gesundheit health insurance with ICD I70.0, I71, I74.0, or I74.1 coding for degenerative aortic disease were in group A.

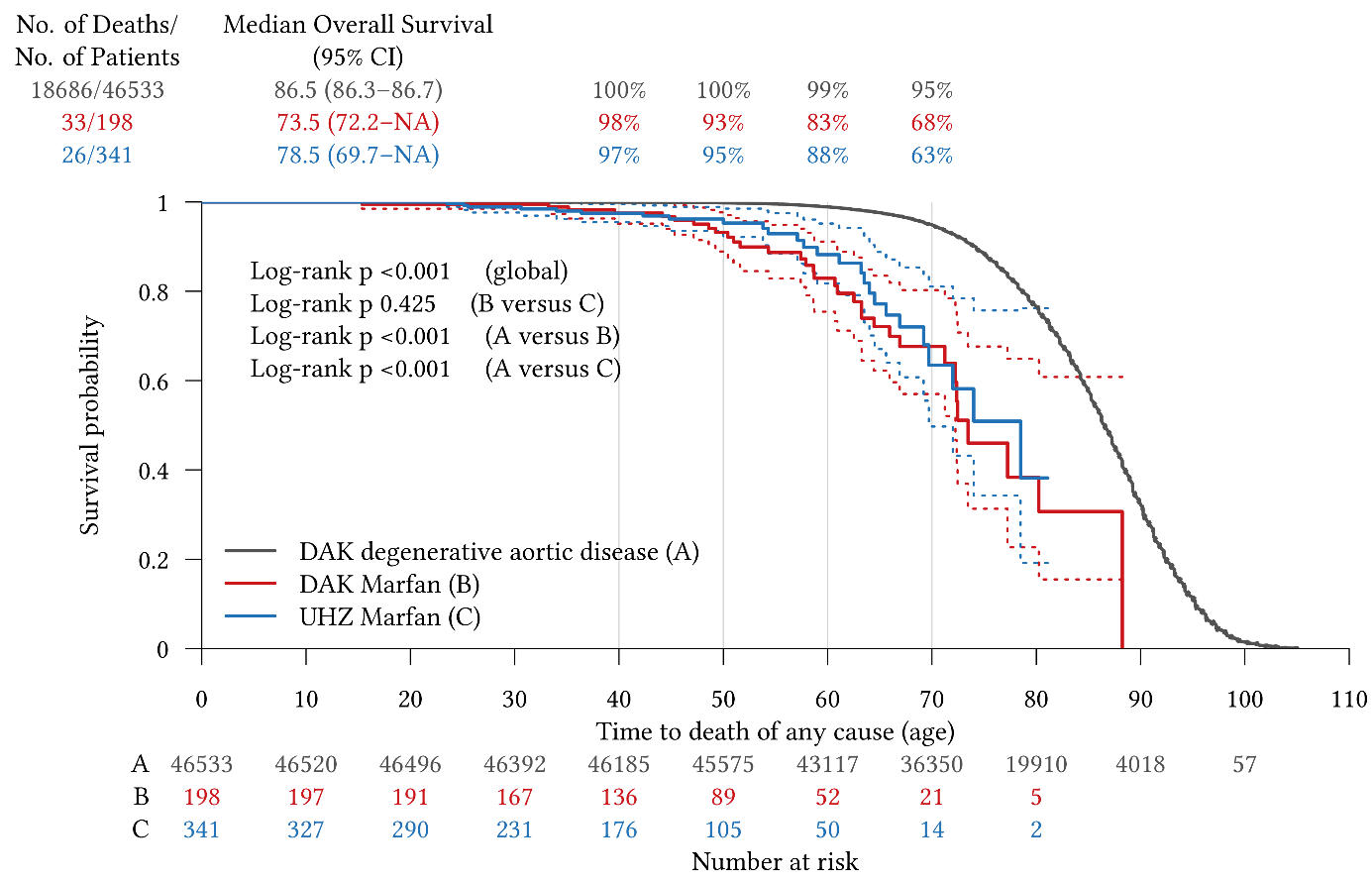

### Figure S8. PRISMA-ScR 2018 flow diagram

The PRISMA extension for scoping review flow diagram for a brief scoping review to qualify the nature of the evidence in the literature on the prognostic impact of time of diagnosis of MFS on aortic events and mortality (PRISMA-ScR) ^5^.

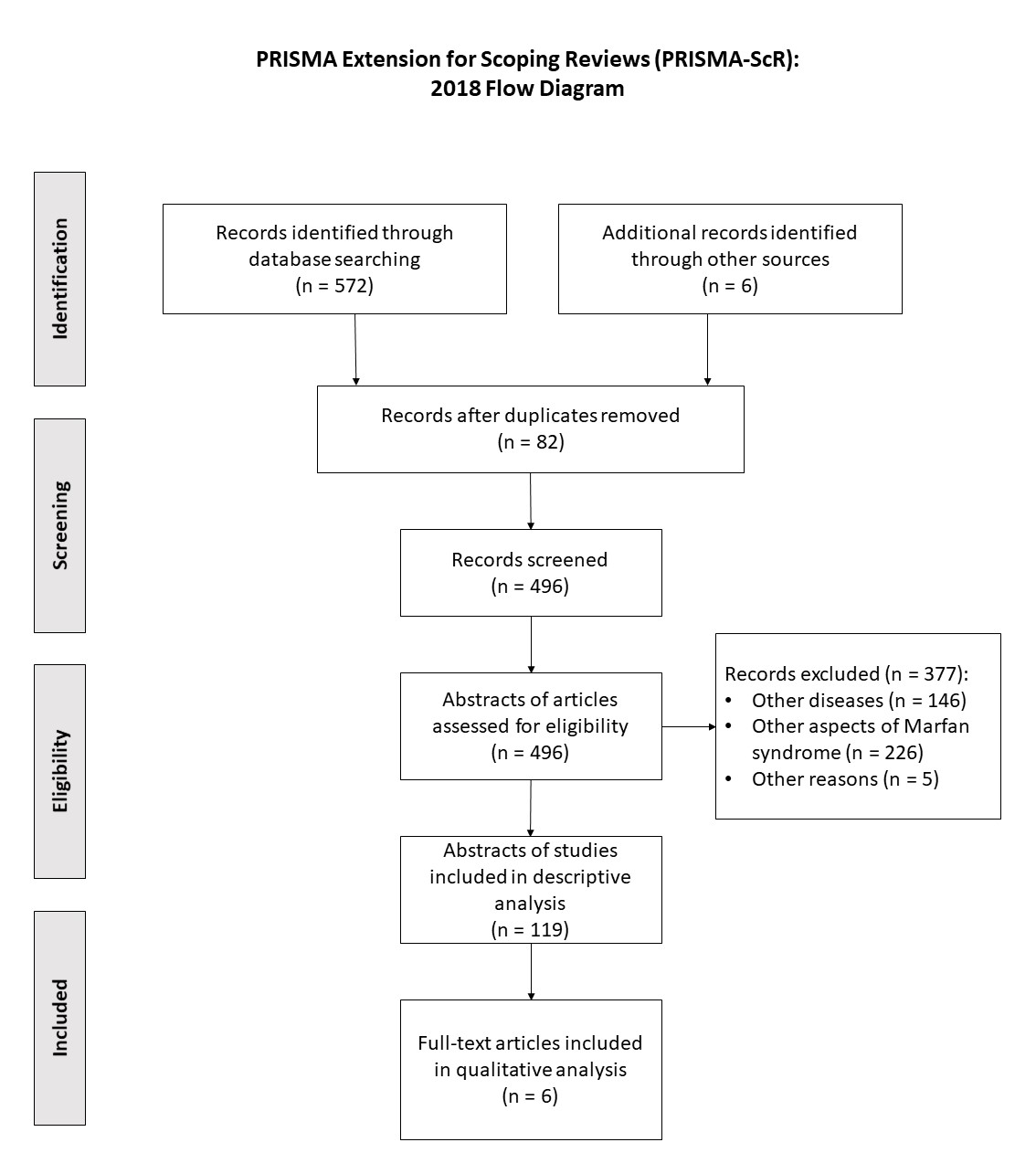

42.	De Cario R, Giannini M, Cassioli G, Kura A, Gori AM, Marcucci R, Nistri S, Pepe G, Giusti B and Sticchi E. Tracking an Elusive Killer: State of the Art of Molecular-Genetic Knowledge and Laboratory Role in Diagnosis and Risk Stratification of Thoracic Aortic Aneurysm and Dissection. *Diagnostics (Basel)*. 2022;12.

43.	Salik I and Rawla P. Marfan Syndrome *StatPearls* Treasure Island (FL): StatPearls Publishing, Copyright © 2022, StatPearls Publishing LLC.; 2022.

97.	Jain E and Pandey RK. Marfan syndrome. *BMJ case reports*. 2013;2013.

98.	O'Brien D, White S, Wilson D, Haworth K and Williams A. Thoracic aortopathies in the military patient. *J R Army Med Corps*. 2015;161:230-6.

99.	Rodríguez-González M, Matamala-Morillo M, Segado-Arenas A, Marín-Iglesias Mdel R and Lechuga-Sancho AM. Chest Pain in Children With Suspected Type I Fibrillinopathy: A Case Report. *Pediatrics*. 2015;136:e1035-8.

100.	Wang Y, Chen S, Wang R, Huang S, Yang M, Liu L and Liu Q. Postmortem diagnosis of Marfan syndrome in a case of sudden death due to aortic rupture: Detection of a novel FBN1 frameshift mutation. *Forensic Sci Int*. 2016;261:e1-4.

101.	AlShehri OA, Almarzouki H, Alharbi BA, Alqahtani M and Allam K. Bilateral posterior crystalline lens dislocations in an otherwise healthy child. *GMS Ophthalmol Cases*. 2017;7:Doc26.
